# Loss of ERG function causes lymphatic vessel malformations and primary lymphoedema

**DOI:** 10.1101/2025.03.10.25323421

**Authors:** D Pirri, H Maude, A Stroobants, K Ogmen, E Sackey, S Dobbins, D Grigoriadis, JC Del Rey Jimenez, DE Al-Ansari, K Frudd, G Saginc, C Peghaire, S Sutanto, T Vujic, R Moy, DOF Gresham, A Manolias, D Nagy, K Riches, J Dean, C Turner, S Ellard, F Elmslie, V Keeley, S Jeffery, P Mortimer, K Gordon, K Paschalaki, T Mäkinen, A Celiz, IR Gould, S Mansour, S Martin-Almedina, AM Randi, I Cebola, P Ostergaard, GM Birdsey

## Abstract

Transcription factor networks are crucial for the regulation of endothelial cell gene expression during vascular development and homeostasis. A recent analysis of 269 rare diseases in 77,539 individuals revealed an association between loss-of-function variants in *ERG*, encoding an ETS transcription factor, with primary lymphoedema. However, the pathogenicity of such variants and possible mechanisms of *ERG*-associated lymphatic vessel dysfunction remains to be established. Here, we have further identified and characterised lymphoedema-associated *ERG* genetic variants, revealing pathogenic mechanisms ranging from differential ERG subcellular localisation to altered DNA-binding and impaired transactivation. We confirm a role for ERG in regulating lymphangiogenesis using *in vitro* assays and a lymphatic endothelium-specific *Erg* deletion mouse model. Furthermore, we characterise the transcriptional and epigenomic landscape of human dermal lymphatic endothelial cells, identifying a unique role for ERG in regulating lymphatic gene programs, including the establishment of cooperative TF networks with PROX1 and GATA2. Our studies identify ERG as a master regulator of lymphatic endothelial cell transcriptional networks and uncover the mechanisms that underpin a novel causative gene for primary lymphoedema.

## Introduction

Blood and lymphatic vessels form interconnected networks that are essential for transport of fluids, gases, macromolecules, and cells. Both vessels are lined by endothelial cells (EC), and although angiogenesis and lymphangiogenesis share common molecular and cellular processes, there is extensive specialisation underlying the unique functions of these two vascular systems^1^, reflected in specific transcriptional and epigenomic features^2,3^. Crucial to lymphangiogenesis and lymphatic EC (LEC) lineage specification and maintenance is the expression of prospero-related homeobox-1 (PROX1), a master regulator of the LEC transcriptional program^1,4^. Primary lymphoedema (PL) is a rare chronically debilitating disease involving lymphatic dysfunction with disturbed tissue fluid balance, which affects about 130,000 people in the UK, of which 28,000 are estimated to have primary lymphoedema (PL)^5^. Under the umbrella of primary lymphatic anomalies^6^, PL is a heterogenic condition with highly variable clinical manifestations, in part due to the diversity of causal genetic mechanisms.

So far, rare variants in ∼40 genes have been reported to cause PL^7^. Several of these genes encode transcription factors (TFs) essential for the development and function of the lymphatic system^6–8^, each leading to specific PL presentations. Examples include *GATA2*-deficiency (Emberger) Syndrome^9–11^, *FOXC2*-associated lymphoedema distichiasis syndrome^12^ and *SOX18*-associated hypotrichosis-lymphoedema-telangiectasia-renal defect syndrome^13,14^. However, approximately 75% of PL patients still lack a genetic diagnosis^6^.

The 100,000 Genomes Project (100KGP) was launched to fulfil the diagnostic gap for rare diseases such as PL, combining genomic sequencing with phenotypic data to identify novel disease-causing variants^15^. We have recently employed Bayesian genetic association tests on whole genome sequencing (WGS) data from the Rare Diseases Main Programme of the 100KGP, identifying 260 associations between genes and rare disease classes, including 19 novel associations^16^. Among these, we identified a novel association between PL and loss-of-function coding variants in the Erythroblast Transformation Specific (ETS)-family TF encoding gene *ERG*^16^. On the other hand, common variants in the *ERG* locus have been associated with susceptibility for varicose veins^17^, a trait that can be present in patients with lymphatic dysfunction. Despite ERG being one of the most highly expressed ETS TFs in blood ECs, where it acts as an essential master regulator of endothelial lineage specification, maintenance and homeostasis^18–23^, there is a lack of knowledge concerning the role of ERG in lymphatic endothelium and the molecular mechanisms of ERG-associated PL remain unclear.

In this study, we uncover the different pathogenic mechanisms through which sequence variants in *ERG* cause PL. Furthermore, we reveal the central role of ERG in lymphatic transcriptional networks and confirm its key function in mammalian lymphangiogenesis.

## Results

### Whole genome sequencing identifies candidate genetic variants in ERG in individuals with primary lymphoedema

In humans, *ERG* is predicted to be intolerant to sequence variation with a LoFtool^24^ score of 0.07 and a LOEUF^25^ score of 0.23. We have previously associated truncating loss-of-function variants in *ERG* with PL^16^. These *ERG* variants were identified using BeviMed^26^, a Bayesian association test that enables prediction of pathogenic variants in rare diseases, but detailed investigations of the contribution of *ERG* genetic variants to PL have not been conducted.

We also queried the WGS data of 100KGP participants with a “Primary Lymphoedema” label for rare missense variants (MAF <0.01%) within the coding regions of the *ERG* gene and identified two novel missense variants in the region close to the ETS DNA-binding domain (**Fig. 1a, Extended Data Table 1**).

**Figure 1.**
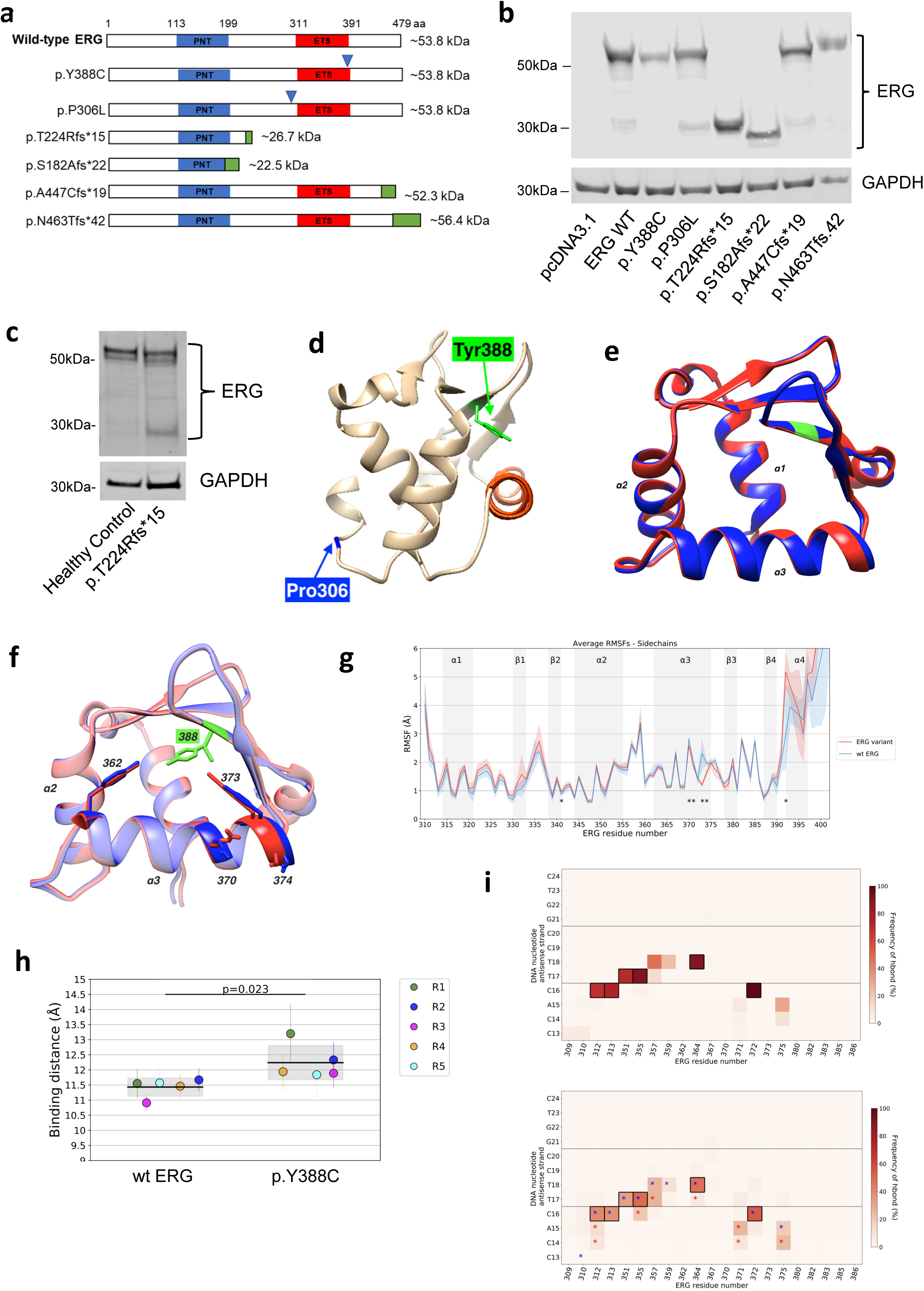
Structural analysis of *ERG* variants associated with primary lymphoedema. **a)** Schematic showing the predicted effects of each variant on the ERG protein product in comparison to the ERG full length canonical polypeptide sequence. The frameshift variants lead to the predicted incorporation of aberrant amino acids (green) downstream of their position and use of alternative stop codons. Predicted size of ERG protein is shown in kDa. PNT, pointed domain (blue); ETS, Erythroblast Transformation Specific DNA-binding domain (red); blue arrows show position of missense variants; aa, amino acids. **b)** Immunoblotting of protein lysates from HEK293 cells transfected with mammalian expression plasmid pcDNA3.1 encoding ERG wild type (WT) or specific ERG variants, as indicated. Membrane is labelled for ERG and GAPDH. Molecular markers are shown on the left. **c)** Representative immunoblot showing ERG molecular weight in ECFC protein lysates from healthy control and ECFC from ERG p.T224Rfs*15, labelled with antibodies to ERG and GAPDH. **d)** Structural representations of the ETS binding domain of wild type ERG derived from the AlphaFold structure (AF-P11308), highlighting the location of two ERG variant sites, Tyr388 (green) and Pro306 (blue), with respect to the DNA-binding helix (orange). **e)** Overlay of the backbone structures of the ETS domain of wild type ERG (blue) and the ERG p.Y388C variant (red), highlighting position of residue Tyr388 (green). **f)** Visual comparison of the average sidechain positions in the α3 DNA-binding helix of ERG WT (blue) and the ERG p.Y388C variant (red). The variant site, residue Tyr388 is shown (green). **g)** Comparison of the sidechain RMSF for each residue of ERG WT (blue) and the p.Y388C variant (red). Significant differences (p <0.05) are marked with an asterisk. **h)** Average DNA binding distance of ERG WT and the p.Y388C variant. The circles represent the average values obtained for one repeat, the black line represents the average value for all repeats and the grey box represents the standard deviation of the repeats. N=5. **i)** Frequency of hydrogen bonds established between residues of ERG WT and DNA nucleotides in the antisense strand (top); frequency of hydrogen bonds established between residues of the ERG p.Y388C variant and DNA nucleotides in the antisense strand (bottom). The boxes represent amino acid-DNA interactions. Asterisks represent a significant difference between the ERG WT and the variant (p<0.05); blue asterisks represent a more frequent interaction in the ERG WT model, red asterisks represent a more frequent interaction in the ERG variant.

MetaDome analysis^27^ indicated that all six variants are in regions intolerant to sequence changes, supporting their potential pathogenicity (**Extended Data Fig. 1a**). The presence of the variants in probands and respective parents was confirmed through inspection of WGS data using Integrative Genomics Viewer (IGV) and, where DNA was available, through Sanger sequencing and co-segregation analysis (**Extended Data Fig. 1b,c**).

Transient overexpression of the six *ERG* variants in HEK293 cells, in which ERG is naturally absent, confirmed their predicted molecular weights (**Fig. 1b**). This was further validated for variant p.T224Rfs*15 using protein lysates of endothelial colony forming cells (ECFCs) from a PL patient (F1:II.1) heterozygous for the variant in which, as expected, immunoblotting revealed two bands for ERG, one corresponding to the full length polypeptide (∼55 kDa) and a lower band (∼27 kDa) corresponding to the expected size for the allele carrying the p.T224Rfs*15 variant (**Fig. 1c**).

### Structural analysis of ERG missense variant Y388C

We hypothesized that *ERG* variants affecting residues within or in the vicinity of the ETS domain, such as p.Y388C and p.P306L, may interfere with ERG’s ability to bind DNA. A previously published crystal structure of the ERG ETS domain^28^ revealed that the Tyr388 residue is near to the DNA-binding helix of ERG, whilst the Pro306 is located further away (**Fig. 1d, Extended Data Fig. 2a**).

Using *in silico* all-atom molecular dynamics, we simulated the structural changes that occur in ERG when carrying the variant p.Y388C. The equilibrated simulations revealed no major structural differences between the two protein sequences (**Fig. 1e, Extended Data Fig. 2b**). However, p.Y388C was predicted to reduce sidechain flexibility in the α3 helix DNA-binding region, as shown by significantly lower root-mean-square-fluctuations (RMSF) values and increased hydrogen bond frequency between residues 370 and 374 (47% to 76%, *p* = 0.001) and between residues 373 and 369 (23% to 44%, *p* = 0.006) (**Fig. 1f,g, Extended Data Table 2**). The p.Y388C variant may also reposition the sidechain of Tyr373 towards the inside of the ETS domain, potentially filling the missing space resulting from the shorter cysteine sidechain in the p.Y388C variant (**Fig. 1f, Extended Data Fig. 2c**).

We then modelled the impact of p.Y388C on ERG binding to the sense and antisense strands of DNA containing conserved ETS motifs. Even though the ETS domain remained bound to DNA throughout the simulations, the tightness of the binding, measured by the average binding distance between the DNA-binding α3 helix and DNA, was significantly higher for the p.Y388C variant (**Fig. 1h**). Further supporting this, p.Y388C showed a significant reduction in hydrogen bonding frequency with the antisense strand of DNA for most contacts identified through crystallography (**Fig. 1i, Extended Data Fig. 2d**). Altogether, the molecular dynamics analyses suggest a molecular mechanism for p.Y388C through structural differences, altered hydrogen bonding and increased rigidity within the α3 DNA-binding helix, which in turn may kinetically disfavour DNA binding and reduce ERG’s regulatory activity.

### Functional variant analysis

To assess the ability of ERG variants to affect the protein’s activity as a transcriptional regulator, we performed immunostaining of HEK293 cells and human dermal lymphatic EC (HDLEC) transiently overexpressing the ERG variants to reveal their sub-cellular localisation (**Fig. 2a, Extended Data Fig. 3a,b**) and luciferase reporter assays using a previously characterised ERG target sequence in the promoter of the thrombomodulin gene (*THBD*)^23^ (**Fig. 2b**). These experiments revealed that variants p.T224Rfs*15 and p.S182Afs*22 completely abrogate ERG’s transactivation ability, which is consistent with their retention in the cytoplasm. The variant p.N463Tfs*42 showed a partial impairment in translocating to the nucleus, concordant with the observed intermediate effect on trans-activation, whilst p.Y388C did not impact translocation to the nucleus but dramatically impaired the ability of ERG to drive promoter activity. The ERG variants p.P306L and p.A447Cfs*19 both showed strong nuclear staining and drove promoter activity to a similar extent as the wild type control.

**Figure 2.**
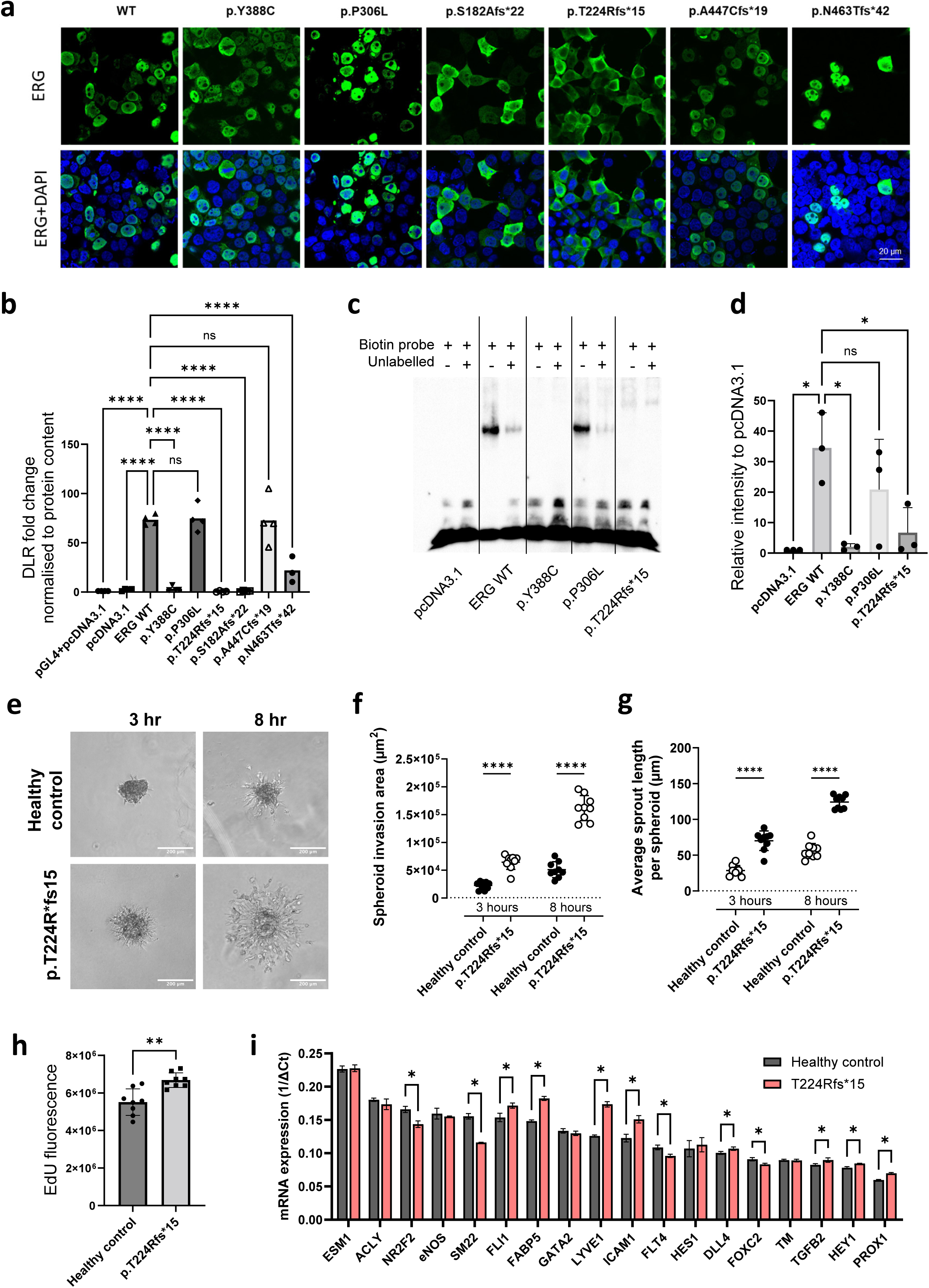
Functional characterisation of *ERG* variants. **a)** Immunofluorescence microscopy of HEK293 cells transfected with mammalian expression plasmid pcDNA3.1 encoding ERG WT or specific ERG variants, as indicated. Cells are labelled for ERG (green) and DAPI (blue). Scale bar, 20 µm. **b)** Plasmids encoding ERG WT or ERG variants were co-transfected into HEK293 cells with luciferase reporter plasmids containing the promoter sequence from the *THBD* gene^23^. Luciferase activity was measured using a Dual-Luciferase Reporter Assay; n=4, One-way ANOVA, Dunnett’s multiple comparison test. **c)** Representative image showing the DNA-binding ability of ERG WT and selected ERG variants using EMSA. ERG DNA binding was imaged using a biotinylated probe containing an ERG target sequence^29^. The DNA-binding efficiency was assessed by adding unlabelled DNA probes. **d)** Quantification of DNA-binding of ERG WT and ERG variants relative to empty vector control (pcDNA3.1); n=4, one-way ANOVA, Dunnett’s multiple comparison test. **e)** Representative light microscopy images of ECFC spheroids from healthy control and ERG p.T224Rfs*15 embedded in 6% gelatin-tetrazine/norbornene hydrogels for 3 & 8 hr. Quantification of spheroid sprouting assay showing spheroid invasion area **(f)** and average sprout length **(g)** per spheroid counted; n=9 spheroids per sample/timepoint, two-way ANOVA, Sidak’s multiple comparison test. **h)** Proliferation assay showing Edu incorporation into donor ECFCs; n=8 technical repeats, unpaired t-test. **i)** Expression of selected genes was determined by RT-qPCR from ECFC RNA lysates. Gene expression was expressed as 1/ΔCt (normalised to *GAPDH*); n=3 technical repeats, unpaired t-test. All data are expressed as mean +/-SD. ns P > 0.05, * P < 0.05, ** P < 0.01, *** P <0.001, **** P < 0.0001.

Electrophoretic mobility shift assays (EMSA) using cell lysates from HEK293 cells overexpressing ERG and a biotinylated oligonucleotide probe, previously shown to be bound by ERG^29^, revealed that the missense variant p.Y388C impaired ERG’s ability to bind DNA to a similar extent as p.T224Rfs*15 (**Fig. 2c,d, Extended data Fig. 3c**), which lacks the entire ETS domain. These results are in line with the molecular dynamics structural analysis of variant p.Y388C, suggesting loss of ERG activity due to structural changes in the ETS DNA-binding domain. In contrast, p.P306L does not alter ERG’s affinity for the probe sequence (**Fig. 2c,d**).

**Figure 3.**
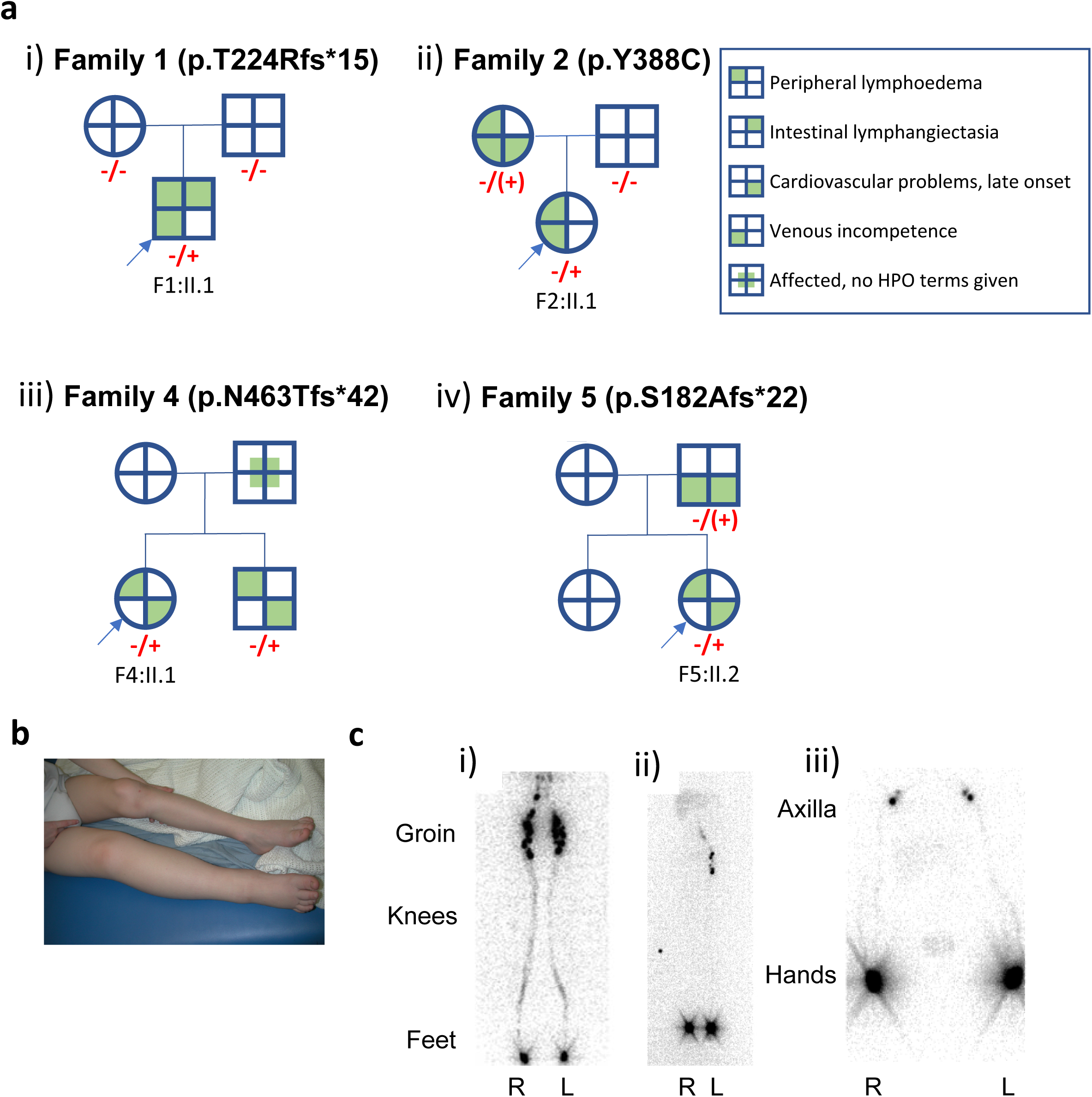
Clinical presentation of ERG-associated primary lymphoedema. **a)** Pedigrees for ERG-associated primary lymphoedema cases. **(i)** Patient ID F1:II.1 (p.T224Rfs*15), **(ii)** F2:II.1 (p.Y388C), **(iii)** F4:II.1 (p.N463Tfs*42), **(iv)** F5:II.2 (p.S182Afs*22). Affected individuals are indicated with filled circles (female) and squares (male). Proband indicated with arrow. Genotypes identified through WGS in the 100,000 Genomes Project are indicated with -/+ (heterozygous for the ERG variant) or -/- (homozygous for the reference allele). Genotypes also confirmed by Sanger sequencing are in red. Notice the mother in (ii) and the father in (iv) are mosaic for the alternative allele indicated with parentheses. **b)** Bilateral lower limb lymphoedema in F1:II.1 (p.T224Rfs*15); right leg worse than left. **c)** Lymphoscintigraphy images at 2 hrs post injection of Technitium-99m. **(i)** In a healthy control, uptake of tracer is observed throughout the lymphatic system and inguinal lymph nodes of both lower limbs. **(ii)** F2:II.1 (p.Y388C) had markedly impaired lymphatic drainage in the left lower limb with reduced uptake of tracer in the left inguinal lymph nodes after 2 hrs. A functional aplasia, where there is no uptake of tracer within the inguinal lymph nodes after 2 hours, was revealed in right lower limb, suggesting the initial lymphatic capillaries fail to absorb and transport tracer, and thus lymphatic fluid, proximally. **(iii)** Although F2:II.1 clinically only presented with swelling of the left hand, her lymphoscintigraphy of the upper limbs showed a bilaterally reduced uptake of tracer (confirmed on quantification data) but via anatomically normal tracts.

ECFCs isolated from a patient heterozygous for the loss-of-function variant p.T224Rfs*15 showed altered morphology compared to sex and age matched healthy control ECFCs, with the patient-derived cells exhibiting a more activated shape (**Extended Data Fig. 3d**). Compared to the control, ECFCs expressing this loss-of-function allele demonstrated an abnormal angiogenic response when grown on Matrigel (**Extended Data Fig. 3e**) or embedded as spheroids in gelatin hydrogel (**Fig. 2e-g, Extended Data Fig. 3f-h**). In line with their altered angiogenic phenotype, Edu incorporation showed an increase in proliferation in ECFCs with the variant p.T224Rfs*15, compared to the control (**Fig. 2h**). RT-qPCR analysis of selected endothelial genes from ECFC RNA lysates suggested that variant p.T224Rfs*15 had a disrupting effect on endothelial homeostasis, with increased expression of *ICAM1* and *TGFB2* (**Fig. 2i**). We also observed dysregulation of lymphatic-associated gene expression, namely an upregulation of *PROX1* and *LYVE1* but a decrease in *FLT4* and *FOXC2* (**Fig. 2i**).

### The phenotype of ERG-associated primary lymphoedema

Following the American College of Medical Genetics and Genomics (ACMG) guidelines^30^, the above experiments revealed that four of the six *ERG* variants identified in 100KGP PL patients are pathogenic (p.T224Rfs*15) or likely pathogenic (p.Y388C, p.N463Tfs*42, and p.S182Afs*22) (**Table 1**). These variants were observed across seven affected individuals from four families, with a family history of lymphoedema consistent with autosomal dominant inheritance in three families (**Fig. 3a**). Interrogation of the 100KGP cohort did not reveal additional non-symptomatic carriers of any of these variants. Furthermore, the seven affected individuals were not carriers of variants affecting other PL genes (see Methods).

**Table 1.** Summary of functional analyses of *ERG* variants and pathogenic classification. Results from the various tests carried out for each variant are summarised here. The strength of the evidence supported a PS3 score for the F1 variant, PS3_moderate for F4 and F5 variants, and PS3_supportive for F2, providing sufficient evidence to classify these variants as likely pathogenic and pathogenic. There was not enough evidence to support the pathogenic effect in F3 and F6, which remain variants of uncertain significance (VUS). Abbreviations: PS3, a strong criterion which can also be used at a moderate and supportive level under the ACMG/AMP sequence variant interpretation framework^91^ used to evaluate well-established in vitro or in vivo functional studies that support a damaging effect on the gene or gene product. *The expected size of the truncated protein if nonsense-mediated decay is not 100% efficient.

| Variant | c.671_672del<br>p.(T224Rfs*15) | c.1163A>G<br>p.(Y388C) | c.917C>T<br>p.(P306L) | c.1386del<br>p.(N463Tfs*42) | c.543del<br>p.(S182Afs*22) | c.1338dup<br>p.(A447Cfs*19) |
| --- | --- | --- | --- | --- | --- | --- |
| Family | F1 | F2 | F3 | F4 | F5 | F6 |
| Size of ERG protein | 237 aa,<br>~26.7 kDa* | 479 aa,<br>~53.8 kDa | 479 aa,<br>~53.8 kDa | 503 aa,<br>~56.4 kDa | 202 aa,<br>~22.5 kDa* | 464 aa,<br>~52.3 kDa |
| Protein modelling | n/a | Pathogenic | n/a | n/a | n/a | n/a |
| Subcellular localisation | Cytoplasm | Nuclear | Nuclear | Partial<br>(nuclear+<br>cytoplasm) | Cytoplasm | Partial (nuclear+<br>cytoplasm) |
| Trans-activation assay | No activity | No activity | WT levels | Low activity | No activity | WT levels |
| EMSA | No DNA binding | No DNA binding | Normal | n/a | n/a | n/a |
| Patient-derived ECFCs | Increased proliferation, disrupted endothelial homeostasis. | n/a | n/a | n/a | n/a | n/a |
| Functional assessment | Pathogenic | Pathogenic | Benign | Likely pathogenic | Pathogenic | Benign |
| PS3 | PS3 | PS3_Supportive | PS3_Supportive | PS3_Moderate | PS3_Moderate | contradictory evidence |
| ACMG criteria | PVS1_strong, PM2, PS3, PP4 | PM2, PP3, PP2, PS3_Supportive PM1_Supportive | PM2, PP2, BP4, PS3_Supportive | PM4, PM2, PS3_Moderate | PVS1_strong, PM2, PS3_Moderate | PVS1_Moderate, PM2 |
| Pathogenicity points | 10 | 6 | 2 | 6 | 8 | 4 |
| Classification | Pathogenic | Likely Pathogenic | VUS | Likely Pathogenic | Likely Pathogenic | VUS |

Six out of seven affected individuals presented with persistent peripheral lymphoedema, mainly in the lower limbs **(Fig. 3b, Extended Data Table 3)**. Three individuals had possible swelling of the upper limbs, but it was milder and sometimes intermittent. Three reported recurrent cellulitis, and one had persistent warts. These complications are frequently observed in lymphoedema patients and thus unlikely to be a problem specifically associated with *ERG* dysfunction. Four affected individuals also had varicose veins or venous incompetence identified by venous duplex. Given the previously reported association of common variants in the *ERG* locus with varicose veins in women^17^, it is possible that this observation reveals a consequence of ERG dysfunction. However, varicose veins are common among PL patients with other genetic aetiologies and thus this feature is not specific to *ERG*-associated PL. Clinical findings in five cases include acquired cardiovascular problems, possibly resulting from hyperlipidaemia (n=2) and obesity (n=3), and lentigines (n=2). One individual (F1:II.1), with a *de novo*, heterozygous variant in *ERG* (p.T224Rfs*15), presented with a progressive generalised lymphatic dysplasia, which in addition to bilateral peripheral lymphoedema, also caused genital oedema with lymphorrhea and intestinal lymphangiectasia (protein losing enteropathy). Lymphoscintigraphy of all four limbs in one patient (F2:II.1; p.Y388C), revealed markedly impaired lymphatic drainage in both lower and upper limbs, compared to a healthy control (**Fig. 3c**). The lower limb lymphoscintigraphy resembled the functional aplasia observed in Milroy patients^31^.

### ERG regulates lymphatic vessel formation

The human genetics findings described above (**Figs. 1-3**), suggest ERG to have a predominant role in lymphatic function and morphogenesis. *In vitro* functional assays were therefore carried out to determine the extent to which ERG controls lymphangiogenesis. Following siRNA inhibition of *ERG in vitro*, HDLEC were less able to form tube-like networks in a 2D Matrigel assay (**Extended Data Fig. 4a,b**), and showed reduced sprouting when HDLEC spheroids were embedded in modified gelatin hydrogel under basal conditions or in the presence of VEGF-C (**Fig. 4a,b**). These results suggested ERG to regulate HDLEC proliferation, which was confirmed by a ∼50% reduction in Edu incorporation following *ERG* inhibition (**Fig. 4c**).

**Figure 4.**
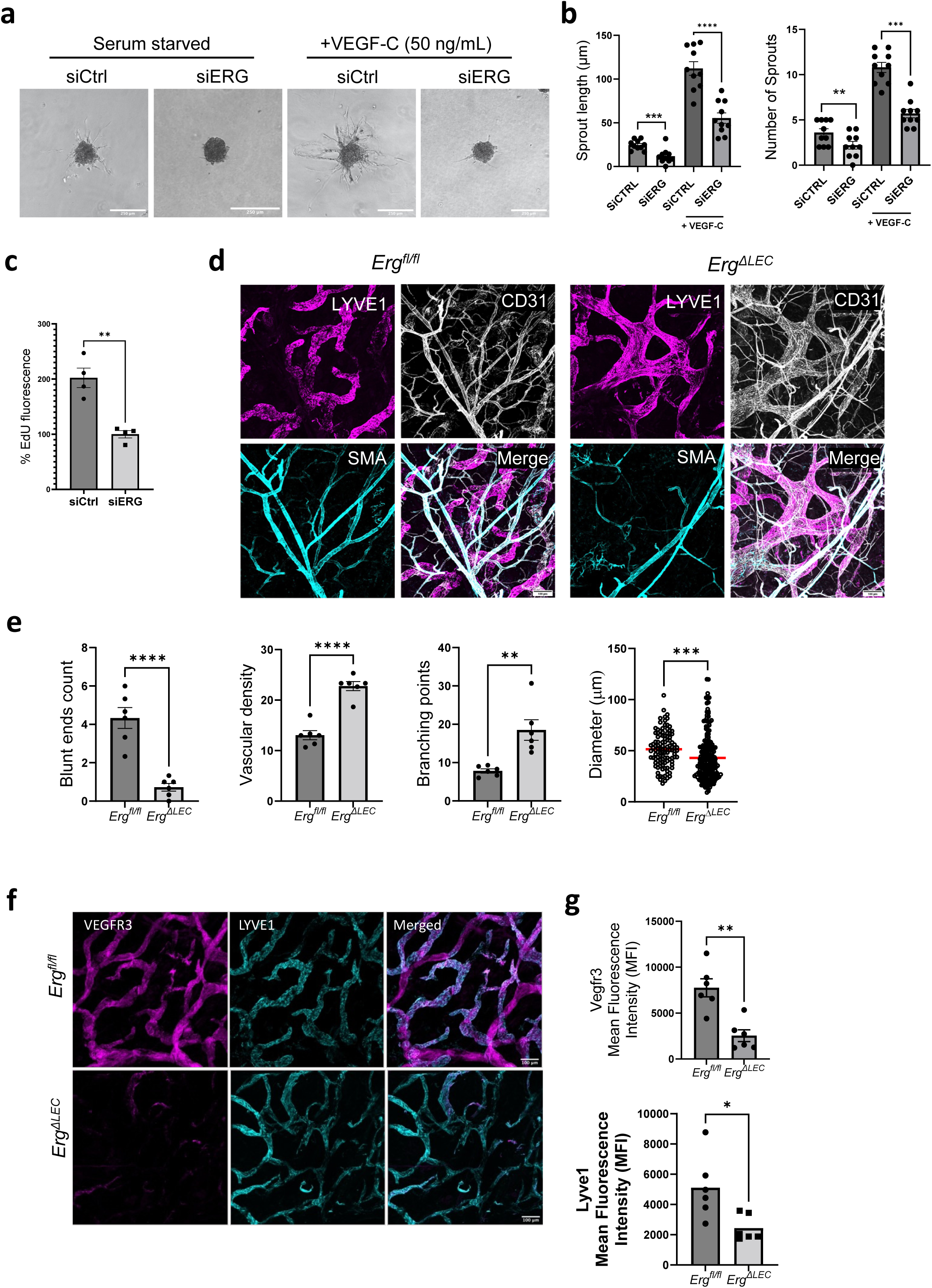
ERG is required for lymphatic vessel formation. **a)** Transmitted light microscopy images showing HDLEC spheroids transfected with non-specific siRNA (siCtrl) or siRNA to ERG (siERG) and embedded in 6% gelatin-tetrazine/norbornene hydrogels in the presence or absence of VEGF-C (50 ng/ml) for 24 hr, as indicated. Scale bar, 250 µm. **b)** Quantification of sprout length and number of sprouts, n = 3, unpaired t-test. **c)** Proliferation assay showing Edu incorporation into HDLEC transfected with siRNA for 48 hr and cultured in EBM2 media containing 1% (v/v) foetal bovine serum, n = 4, unpaired t-test. **d)** Immunofluorescent microscopy of ear dermis from *Erg^fl/fl^* and *Erg^ΔLEC^* mice stained with LYVE1 (magenta), CD31 (grey) and smooth muscle actin (SMA, cyan). Scale bar, 100 µm. **e)** Quantification of vascular structures: numbers of LYVE1+ blunt-ended vessels, numbers of LYVE1+ vessels per field of view (vascular density), numbers of vessel branching points, diameter of LYVE+ vessels; n=6, unpaired t-test. **f)** Immunofluorescent microscopy of ear dermis from *Erg^fl/fl^* and *Erg^ΔLEC^* mice stained for Vegfr3 (magenta) and Lyve1 (cyan). Scale bar, 100 µm. **g)** Quantification of Vegfr3 mean fluorescent intensity (MFI) within the Lyve1+ lymphatic vessels, n=6, unpaired t-test. Quantification of Lyve1 MFI in mouse ear dermis, n=6, unpaired t-test. * P < 0.1, ** P < 0.01, *** P < 0.001, **** P < 0.0001.

To establish the role of ERG in lymphangiogenesis *in vivo*, we used an inducible pan-endothelial-specific ERG deletion model (*Cdh5-CreERT2-ERG^fl/fl^*; referred to as *ERG^iECKO^*)^20^. *ERG^iECKO^* and littermate control (*ERG^fl/fl^*) neonatal mice were administered tamoxifen at postnatal days 1-3 (P1-P3) and the dermal lymphatic vasculature of the ear was analysed by *en face* immunofluorescence microscopy 21 days after birth (P21) (**Extended Data Fig. 4d**). In the absence of Erg, Lyve1^+^ lymphatic vessels appeared malformed, with some vessels showing significant changes in diameter compared to littermate controls (**Extended Data Fig. 4e,f**).

To overcome any potential indirect effects of deleting *Erg* in both blood vessel EC and lymphatic EC, we generated an inducible LEC-specific *Erg* deletion model (*ERG^ΔLEC^*) by crossing *ERG^fl/fl^* mice with the well-characterised LEC-specific Cre recombinase line *Prox1-CreERT2* ^32^. *ERG^ΔLEC^* and *ERG^fl/fl^*neonatal mice were administered tamoxifen at P1-P3 and the dermal lymphatic vasculature of the ear was analysed at P21 (**Extended Data Fig. 4g)**. In the *ERG^ΔLEC^* mice, we confirmed that *Erg* was specifically deleted in the EC of the lymphatic vessels, whilst Erg staining remained detectable in capillaries, arteries, and veins of the blood vasculature (**Extended Data Fig. 4h,i**), with no observable changes in blood vessels of the P21 mouse retina (**Extended Data Fig. 4j**). In the ear skin of *ERG^ΔLEC^* mice, there was a dramatic reduction in the numbers of Lyve1^+^ blunt-ended lymphatic vessels, accompanied by a significant increase in vessel network density and branch points, along with a decrease in mean vessel diameter (**Fig. 4d,e**). Recent analyses of cultured HDLEC suggested that ERG is a crucial regulator of VEGF-C signalling via direct regulation of *VEGFR-3/FLT4* ^33^. In line with this, we observed a significant reduction in Vegfr3 protein levels in dermal lymphatic vessels in the *ERG^ΔLEC^*mice compared to controls (**Fig. 4f,g**). We also detected a small but significant decrease in Lyve1 staining intensity in the *ERG^ΔLEC^* mice compared to controls (**Fig. 4g**).

To prevent lymph backflow, collecting lymphatic vessels contain intraluminal valves. Several key PL-associated genes such as *FOXC2, GATA2, FAT4,* and *EPHB4* are involved in the formation and maintenance of lymphatic valves^10,34–37^. To investigate whether ERG has a role in lymphatic valve formation, we carried out whole-mount immunofluorescence microscopy of P21 mouse ear skin labelled for integrin-α9, expressed in lymphatic valves and a well-established regulator of lymphatic valve development^38^. *ERG^ΔLEC^* mice showed a significant reduction in the numbers of integrin-α9-positive lymphatic valves (**Fig. 5a,b**) with horizontal flattening of the bivalve structures (**Fig. 5c**), reminiscent of the abnormal lymphatic valves in *Itga9*-deficient mice^32^. These observations contrasted with control mice, which exhibited characteristic V-shaped lymphatic valves, labelled strongly for integrin-α9 and Prox1 (**Fig. 5c**). These results suggest that loss of ERG may lead to reduced functionality of the lymphatic valves.

**Figure 5.**
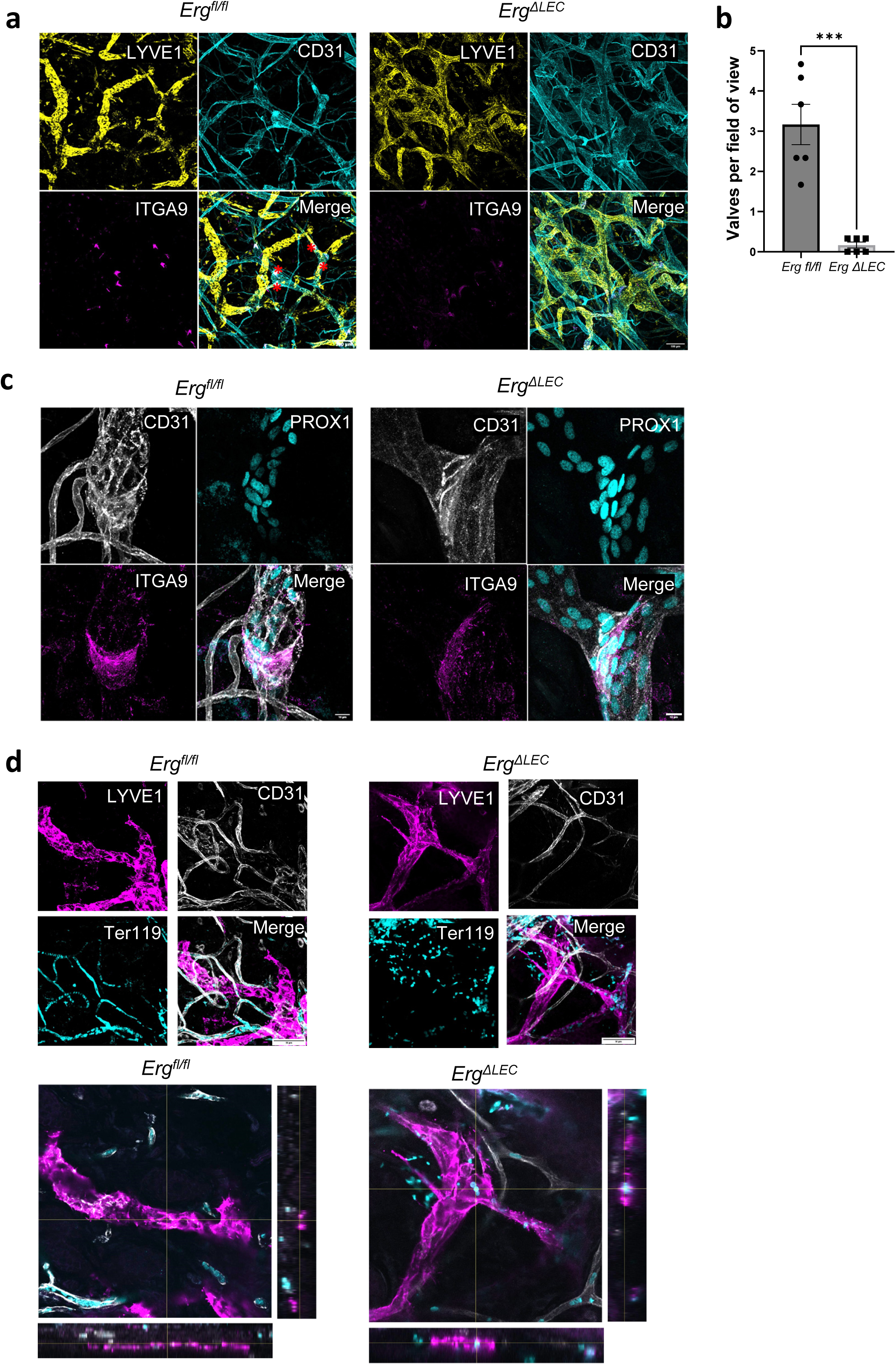
ERG is required for lymphatic valve formation. **a)** Immunofluorescent imaging of ear dermis from *Erg^fl/fl^* and *Erg^ΔLEC^* mice showing lymphatic vessels stained with LYVE1 (yellow), all vascular structures stained with CD31 (cyan), lymphatic valves stained with integrin alpha-9 (ITGA9; magenta). Scale bar, 100 µm. **b)** Quantification of average number of lymphatic valves per field of view; n=6, unpaired t-test. *** P < 0.001. **c)** Higher magnification images of valve structures, as in **a**, labelled for PROX1 (cyan), CD31 (grey), ITGA9 (magenta). Scale bar, 10 µm. **d)** Immunofluorescent imaging of ear dermis from *Erg^fl/fl^*and *Erg^ΔLEC^* mice stained with LYVE1 (magenta), CD31 (grey), and Ter119 (labels red blood cells; cyan). Scale bar, 50 µm; n=5. Representative orthogonal sections below each image show cross-sectional views. Scale bar, 50 µm.

To investigate whether deletion of *Erg* causes defects in blood-lymph separation, we stained mouse ear skin for the red blood cell marker Ter119. As expected, in the *ERG^fl/fl^* mice Ter119-positive red blood cells were exclusively contained within CD31^+^/Lyve1^−^ blood capillaries, whilst Lyve1^+^ lymphatic vessels were devoid of Ter119 staining (**Fig. 5d**). However, in *ERG^ΔLEC^* mice there was an infiltration of Ter119-positive red blood cells within the lymphatic vessels and surrounding dermal tissue (**Fig. 5d**). In summary, ERG is important for the post-natal morphogenesis of an organised dermal lymphatic network and maintaining blood-lymph separation.

### ERG is a major regulator of HDLEC transcriptional landscape

ERG regulates blood vessel endothelial transcriptional networks^21–23^, but its specific role in lymphatic EC transcription networks and chromatin architecture remains largely unexplored. To understand the role of ERG in lymphatic homeostasis, we started by characterising its role in defining the chromatin landscape of lymphatic EC. We carried out assay for transposase-accessible chromatin with high-throughput sequencing (ATAC-seq) and chromatin immunoprecipitation sequencing (ChIP-seq) for H3K27ac in HDLEC (**Fig. 6a, Supplementary Table 1**). Combined with HDLEC Cap Analysis of Gene Expression (CAGE) data from FANTOM6 ^39^, these data were used to create genome-scale HDLEC epigenome annotations, identifying 86,393 *cis*-regulatory elements (CRE) of which 29,764 were defined as active enhancers (**Fig. 6a, Supplementary Table 2**). Many of the identified HDLEC CREs were shared with human umbilical vein EC (HUVEC) (**Extended Data Fig. 5a**). Notwithstanding, we detected 29,910 HDLEC CREs (∼35% of total) that are not accessible in HUVEC (**Extended Data Fig. 5a**). Enhancer clusters, also known as super-enhancers (SE), are important for the regulation of cell type-specific gene expression and of pathways important for defining lineage identity^40^, including endothelial identity as seen in HUVEC^22^. Using the active CRE histone mark H3K27ac, we identified 1,131 SE in HDLEC (**Fig. 6b, Supplementary Table 4**). These included SE associated with genes crucial for lymphatic lineage identity and lymphangiogenesis, including *PROX1, FOXC2, GATA2,* and *FLT4* ^7^. Interestingly we also mapped a HDLEC SE in the *ERG* locus (**Fig. 6b**), supporting this gene’s importance for LEC identity and function.

**Figure 6.**
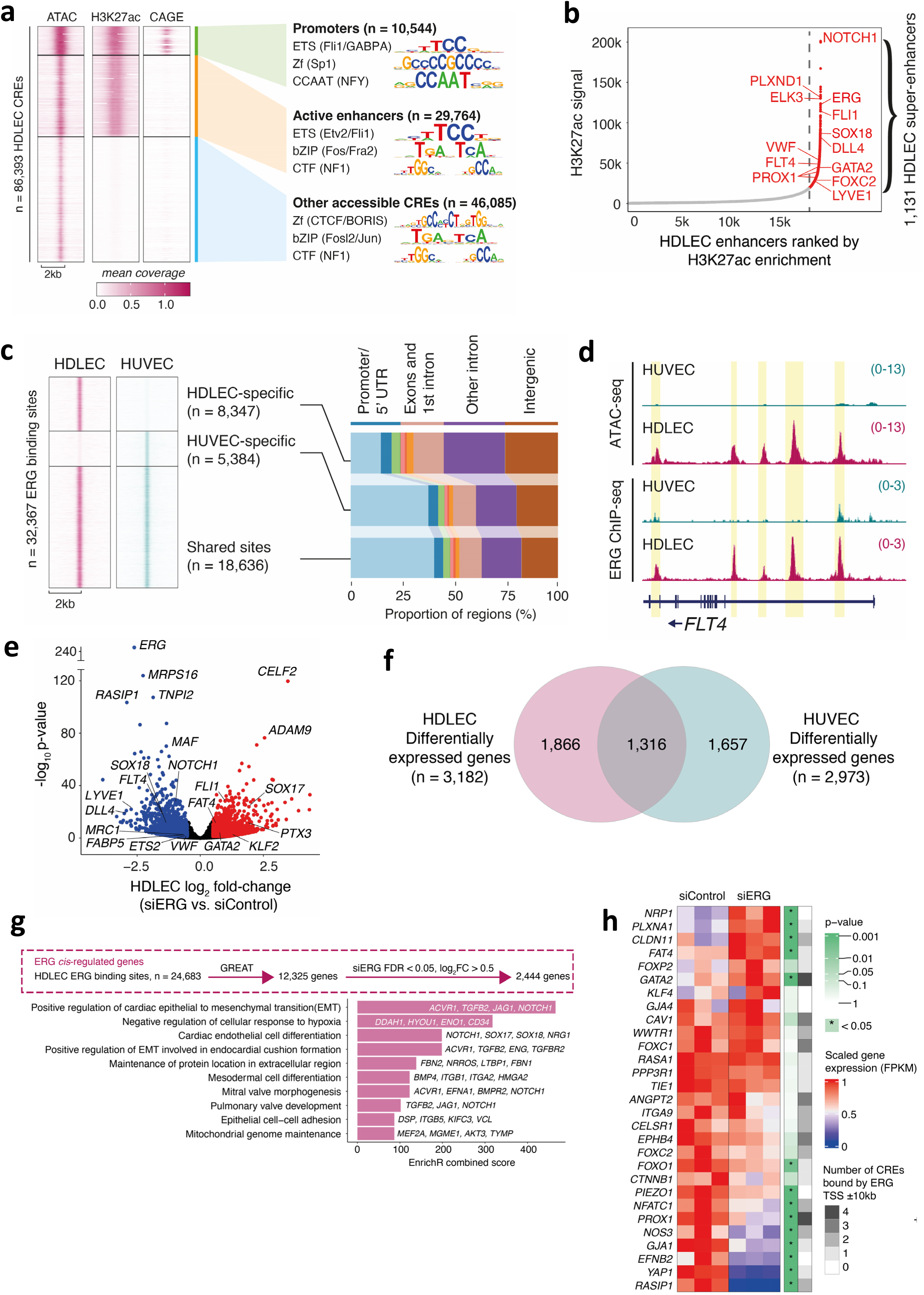
ERG is a master regulator of human LEC transcriptional signature. **a)** HDLEC accessible chromatin regions (ATAC-seq) were classified as promoters or enhancers based on enrichment of H3K27ac and CAGE signal. The heatmaps show HDLEC BPM-normalised coverage of pooled ATAC-seq (n = 2), H3K27ac ChIP-seq (n = 1), and CAGE (n=3) from FANTOM6. The top three families of enriched TF motifs, compared to the genomic background, are shown for the three indicated categories of CREs. **b)** Stitched enhancer clusters are ranked by H3K27ac enrichment, with those to the right of the cutoff (grey dashed line) defined as super-enhancers. Highlighted examples show genes associated with EC and particularly LEC-specialised functions. **c)** Comparison of HDLEC and HUVEC ERG binding profiles, showing the proportions of shared and EC subtype-specific binding sites using a “high-confidence” list of ERG peaks (macs2 q-value < 0.001). The heatmaps show ERG ChIP-seq BPM-normalised coverage (n = 1 per EC sub-type). Bar graphs on the right show the distribution of functional annotations. **d)** Genome browser screenshot presenting HDLEC and HUVEC ATAC-seq and ERG ChIP-seq profiles at the *FLT4* locus (tracks show BPM-normalised coverage). Several regions show ERG binding enriched in HDLEC (yellow boxes). **e)** Inhibition of ERG in HDLEC for 48 hrs leads to widespread changes in gene expression. Blue and red points show genes with significantly decreased or increased expression, respectively, with Benjamini-Hochberg false discovery rate (FDR) adjusted p-value < 0.05 and log2FC < -0.5 or log2FC > 0.5. **f)** Overlap of differentially expressed genes from siERG in HDLEC and HUVEC (adjusted p-value < 0.05 and absolute log2FC > 0.5). **g)** Enrichment of GO biological processes for genes annotated as putative *cis*-regulated targets of all ERG binding sites in HDLEC. **h)** Heatmap showing scaled gene expression (FPKM) of selected LEC valve-associated genes in HDLEC treated with siControl and siERG. The p-value scale (in green) shows the adjusted p-value from DESeq2 comparing siERG to siControl, with significance (non-directional Wilcoxon test, p-value = 0.045) indicated by an asterisk. For each gene, the number of CREs bound by ERG within +/-10 kb of the transcription start site (TSS) are plotted in greyscale.

The strong enrichment for ETS motifs, including ERG, in HDLEC promoters and enhancers (**Fig. 6a, Supplementary Table 3**) led us to postulate that ERG may bind to a large proportion of HDLEC CRE. To address this question, we profiled the global binding landscape of ERG in HDLEC by ChIP-seq (**Fig. 6c, Supplementary Table 5**). This revealed that 50% of promoters and 44% of active enhancers were bound by ERG, supporting the central role of ERG in defining the epigenomic landscape of LEC. When compared to HUVEC^22^, we observed 8,347 HDLEC-specific and 5,384 HUVEC-specific ERG binding sites, whereas 18,636 sites were bound by ERG in both EC sub-types, indicating a large proportion of ERG binding sites to be EC sub-type dependent, including examples at key HDLEC genes such as *FLT4* (**Fig. 6c,d**).

We then carried out RNA-seq on HDLEC and HUVEC treated with siRNA against ERG for 48 hours (**Extended Data Fig. 5b,c**). ERG knockdown in both EC sub-types resulted in global transcriptional changes compared to control cells, with approximately 3,000 differentially expressed genes (DEGs) in each EC sub-type (**Fig. 6e, Extended Data Fig. 5d, Supplementary Table 6**). Most DEGs were specific to either HDLEC or HUVEC (**Fig. 6f, Extended Data Fig. 5e,f)**. Selected putative ERG targets in HDLEC were validated for mRNA and protein levels, confirming significant changes in expression of key genes involved in lymphatic function (**Extended Data Fig. 5g-j**). By integrating the RNA-seq and ERG ChIP-seq datasets from HDLEC, we observed that 75% of DEGs were near at least one ERG binding site, revealing 2,444 direct targets of ERG. Pathway analysis of these ERG-regulated genes showed enrichment in endothelial cell differentiation, valve formation and regulation of cell adhesion (**Fig. 6g**). Cell type-specific CREs tend to regulate genes involved in cell type-specific functions^41^; accordingly, ERG targets with HDLEC-specific ERG binding sites (n=1,452 genes) were enriched in lymphatic endothelial cell differentiation and valve morphogenesis (**Extended Data Fig. 5k**). Gene set enrichment analysis of genes implicated in lymphatic valve formation further confirmed a significant enrichment for differential expression following inhibition of ERG in HDLEC (**Fig. 6h**), in line with our experimental data in *ERG^ΔLEC^* mice (**Fig. 5**), suggesting a role for ERG in regulating the transcriptional programs for formation and/or remodelling of lymphatic valve structures.

### ERG and PROX1 are key partners in LEC TF network

TF cooperativity is an essential mechanism for the appropriate regulation of vascular EC function and homeostasis^2^. Given the observed activity of ERG in HDLEC and its frequent binding to HDLEC CREs (**Fig. 6**), we postulated that ERG may be part of an important LEC transcriptional network. We thus explored the relationship between ERG and four key transcriptional regulators of LEC identity (PROX1, GATA2, NFATC1, and FOXC2) at the chromatin level using ChIP-seq datasets from HDLEC^42^. ERG co-binds with all four LEC TFs at an enhancer previously established to be essential for *PROX1* expression and LEC identity^42^ (**Fig. 7a**). A second example at the *FLT4* locus demonstrates distinct binding patterns, particularly of GATA2 and FOXC2, however there are nearly identical ERG and PROX1 co-occupancy patterns (**Fig. 7b**).

**Figure 7.**
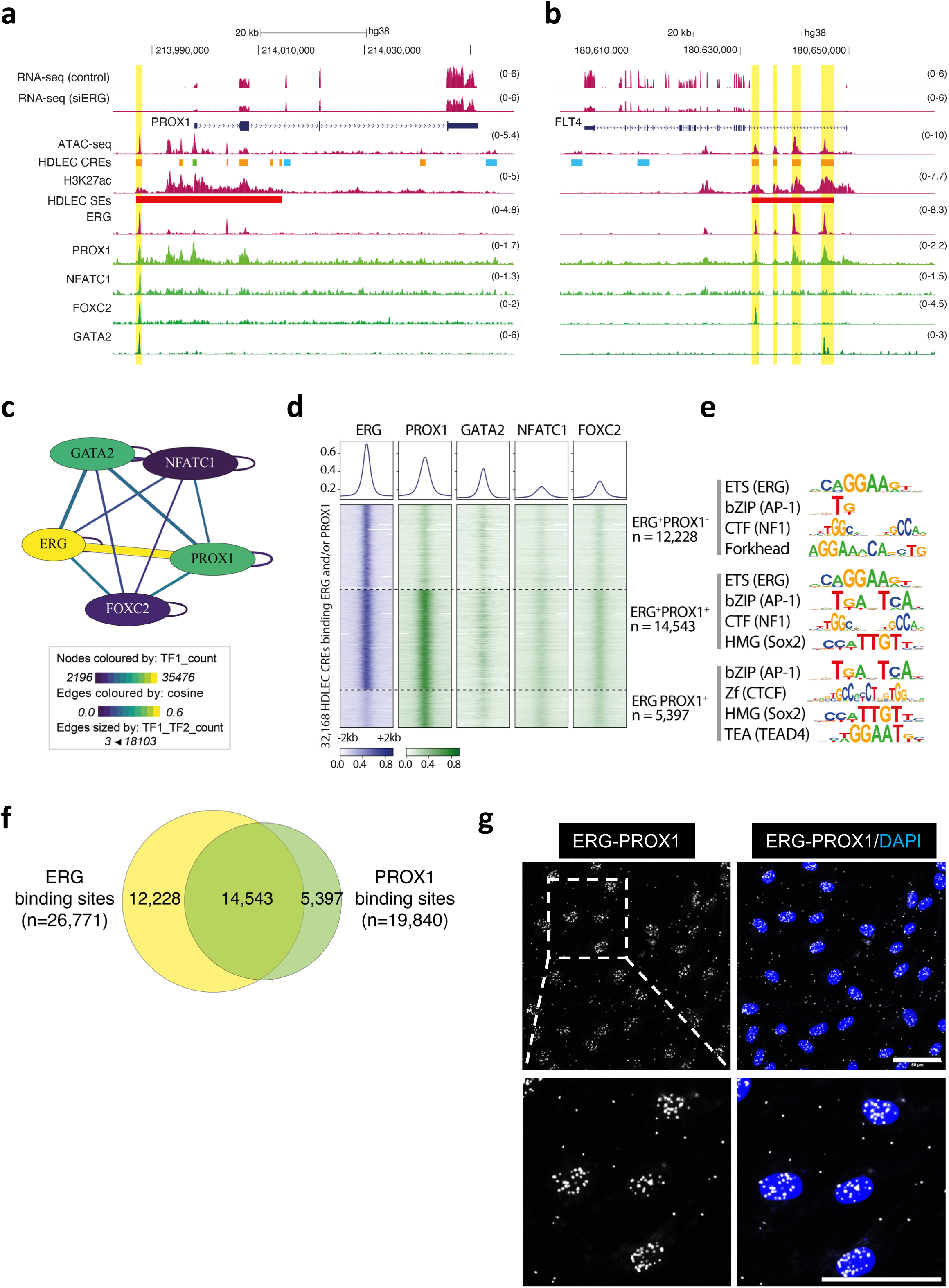
ERG is part of a LEC transcription factor network. Genome browser screenshots for **(a)** *PROX1* and **(b)** *FLT4* loci, showing HDLEC RNA-seq (n = 3/condition), ChIP-seq (H3K27ac and ERG, n = 1), and ATAC-seq (pooled, n = 2) datasets (magenta tracks, generated as part of this study) together with ChIP-seq for key LEC TFs (PROX1, NFATC1, FOXC2 and GATA2; n = 1/TF) (green tracks; data generated by Kazenwadel et al. 2023 ^42^ and re-analysed in-house). The HDLEC CRE track shows the classification of regulatory elements with promoters in green, enhancers in orange, and other accessible elements in blue. HDLEC super-enhancers (SE) are shown as red bars. In **a**, the *PROX1* essential enhancer is highlighted in yellow. In **b**, yellow highlights show CREs co-bound by ERG and PROX1 at the *FLT4* locus. **c)** TF-COMB network map showing the quantification of binding site overlaps for ERG, PROX1, GATA2, NFATC1 and FOXC2. Nodes are coloured by the total number of TF-bound CREs (intersection of HDLEC CREs with ChIP-seq peaks, macs2 q-value < 0.05), while connecting lines (Edges) are coloured by the cosine association score, which represents the frequency of TF co-occurrence at HDLEC CREs. The thickness of the lines reflects the absolute count of TF co-occurrence. **d)** Heatmap of BPM-normalised ChIP-seq coverage (n = 1/TF) at HDLEC CREs bound by ERG (blue) and the other LEC TFs (green), as indicated. CREs are grouped by ERG and/or PROX1 binding. **e)** Top enriched transcription factor motif families for the different classes of HDLEC CRE, based on ERG and PROX1 binding. **f)** Overlap of ERG and PROX1 binding sites in HDLEC. **g)** Representative images of proximity ligation assay (PLA) showing nuclear ERG-PROX1 interactions (white signal) in confluent HDLEC, n=3. Nuclei are identified by DAPI (blue). Scale bar, 50 µm.

These locus-specific observations led us to systematically examine the relationship between the ERG and the other four TFs, based on their occupancy patterns. TF-COMB (Transcription Factor Co-Occurrence using Market Basket)^43^ revealed that (i) ERG is part of a HDLEC TF network, together with PROX1, GATA2, NFATC1, and FOXC2; and (ii) among these five TFs, the global binding profiles of ERG and PROX1 have the highest similarity (**Fig. 7c, Extended Data Fig. 6a**). Consistent with ERG being part of a core LEC TF network, the addition of GATA2, PROX1, FOXC2 and NFATC1 to CREs bound by ERG increased the specificity of target gene expression in HDLEC compared with other EC sub-types, with the highest levels of HDLEC-enriched expression detected for genes associated with CREs bound by all five TF (**Extended Data Fig. 6b-c, Supplementary Table 8**). Moreover, analyses of the overlap between HDLEC CREs and ChIP-seq peaks for the five TFs confirmed the preferential co-occurrence of ERG with PROX1, compared to the other LEC TFs. In total, 37% (n = 32,168/86,393) of HDLEC CREs were bound by either ERG or PROX1 and 73% of PROX1^+^ CREs were also bound by ERG (**Fig. 7d,f, Extended Data Fig. 6d, Supplementary Table 8**). Despite the strong co-occurrence of ERG and PROX1, their genomic distribution showed some differences, with sites bound by PROX1 more likely to be promoters, while ERG sites not bound by PROX1 were more likely to be distal CREs (p-value < 2.2 × 10^−16^, Fisher’s exact test; **Extended Data Fig. 6e**). As expected, ERG was the top enriched TF motif for ERG-bound sites (including those co-bound by PROX1). However, CREs bound by PROX1 but lacking ERG did not show enrichment for ERG or a PROX1-specific motif (**Fig. 7e, Supplementary Table 9**), being instead enriched for bZIP, HMG and TEAD factors, possibly suggesting distinct PROX1 binding partners at this subset of CREs.

TF binding site co-occurrence, based on ChIP-seq profiling, does not necessarily imply functional cooperativity nor simultaneous occupancy by different TFs at the same CREs. We thus sought to investigate if ERG physically interacts with PROX1 or GATA2, the two TFs that showed the greatest degree of co-occurrence with ERG in our *in silico* analyses (**Fig 7c, Extended Data** Fig 6d). Using *in situ* proximity ligation assays (PLA) in HDLEC, we observed that ERG physically interacts with PROX1 and GATA2 (**Fig. 7g, Extended Data Fig. f-h**). Moreover, inhibition of ERG with siRNA specifically affected ERG-PROX1 PLA signal but had no significant effect on the PROX1-GATA2 interaction (**Extended Data Fig. 6g-h**). Together, these data reveal a novel cooperative TF network in LEC, of which ERG and PROX1 appear to be the dominant partners, and uncover molecular mechanisms through which ERG is a causative gene for PL.

## Discussion

We recently described an association between loss-of-function variants in *ERG* and primary lymphoedema (PL)^16^. However, the molecular mechanisms through which ERG transcriptionally regulates LEC gene expression and how this may be disrupted by ERG variants has not been established.

Two additional ERG variants were included in this study and through multiple *in vitro* and *in silico* approaches we observed how some of the variants lost functional activity, concluding that four of the six investigated *ERG* variants are likely pathogenic (LP) or pathogenic (P). The positions of the p.S182Afs*22 and p.T224Rfs*15 variants suggest nonsense-mediated decay (NMD) and haploinsufficiency as an additional disease mechanism. However, the presence of the shorter ERG peptide in ECFCs from a patient heterozygous for p.T224Rfs*15 indicates that at least some of this ERG mRNA transcript evades NMD, and the loss of function phenotype may be due to ERG mislocalization outside of the nucleus.

The affected PL patients carrying heterozygous P/LP *ERG* variants present with late onset peripheral oedema (particularly the lower limbs), varicose veins, and are possibly prone to obesity and hyperlipidaemia, but the numbers are too small to prove an association with these clinical features. Additionally, only one proband had significant systemic involvement with genital lymphorrhea and intestinal lymphangiectasia.

How the ERG variants cause lymphatic disruption is not known, but we found ERG binding at an enhancer region previously shown to be crucial for *Prox1* expression and lymphatic endothelial cell identity^42^. This region was shown to be also bound by GATA2, FOXC2, NFATC1 and PROX1. These were previously identified as essential regulators of the lymphatic valve development, and their loss in mouse models led to lymphatic valves agenesis ^35,42,44^. In line with this, using LEC-specific *Erg* deletion model (*ERG^ΔLEC^*), we observed aberrant lymphatic valves at reduced numbers in the mouse ear dermis. The postnatal loss of *Erg* in mouse LECs also had profound effects on lymphangiogenesis leading to loss of blunt-ended capillaries. Lymphatic network maturation and maintenance is dependent on intact VEGFC–VEGFR3 signalling^45–48^ and ERG could be involved in this, as we also demonstrated ERG as a regulator of VEGFR3 (encoded by *FLT4*) in HDLECs and in the *ERG^ΔLEC^* mice, in agreement with a recent study by Yamashita *et al*.^33^

In conclusion, our study shows there is a unique ERG-dependent LEC transcriptional signature, and we provide evidence for a novel interaction between ERG and core lymphatic TFs, including PROX1 and GATA2. We provide compelling evidence for Erg’s role in mouse lymphatic network morphology and patterning, and that ERG variants can be determinant for primary lymphoedema in patients.

## Methods

### Cell culture

HEK293 cells were cultured in Dulbecco’s Modified Eagle Medium (DMEM; Invitrogen) containing 4.5 g glucose, supplemented with 1% L-glutamine, 1% Penicillin/Streptomycin and 10% Hyclone Fetal Bovine Serum (FBS, South American origin). Single donor human dermal lymphatic endothelial cells (HDLEC from juvenile foreskin, C-12216 [Lot numbers: 470Z021.2, 4397Z007.2, 440Z009.2, 489Z014]; PromoCell, Heidelberg, Germany) were grown in EGM-MV2 media (C-22121; PromoCell). Pooled donor human umbilical vein endothelial cells (HUVEC, C2519A; Lonza) were cultured in complete EGM-2 media (CC-3162; Lonza). HDLEC and HUVEC were cultured on 1% (v/v) gelatin-coated plasticware in a humidified chamber at 37 °C in 5% CO_2_. In some experiments, HDLEC were cultured on 1 ug/mL fibronectin from human plasma (F2006; Sigma-Aldrich). Endothelial cells were transfected with siRNA (20 nM) against ERG exon 6 (siERG, 5ʹ-CAGATCCTACGCTATGGAGTA-3ʹ; Qiagen) or with AllStars Negative Control siRNA (siCtrl, 20 nM; Qiagen) using Lipofectamine RNAiMAX Transfection Reagent (Invitrogen) for 48 hr.

### RNA-seq, ChIP-seq and ATAC-seq sample preparation

For RNA-seq, HUVEC and HDLEC were grown in triplicate on 6-well plates and transfected with 20 nM control or ERG siRNA for 48 hr. Total RNA was extracted from cell lysates using the RNeasy Kit (Qiagen), according to the manufacturer’s instructions. For ChIP-seq, HDLEC were cultured to confluency and then fixed for 15 min at room temperature using 11% formaldehyde (F-8775; Sigma-Aldrich) containing 0.1 M NaCl, 1 mM EDTA and 50 mM HEPES. Fixation was stopped using 2.5 M Glycine solution (G7403; Sigma-Aldrich). Cells were collected by scraping and resuspended in 0.5% Igepal (I-8896; Sigma-Aldrich) in PBS, containing 1 mM phenylmethanesulfonyl fluoride (PMSF, P-7626; Sigma-Aldrich). Chromatin immunoprecipitation was carried out by Active Motif using rabbit anti-ERG antibody (sc-354X; Santa Cruz). For ATAC-seq, HDLEC (1 × 10^5^ cells) were cultured to confluency on gelatin-coated plates, detached using Trypsin-EDTA, and resuspended in 500 µL of cryopreservation solution (50% FBS, 40% growth media, 10% DMSO). Cells were stored frozen at -80 °C. Next generation sequencing for all three assays was carried out by Active Motif using Illumina NextSeq 500. RNA-seq and ATAC-seq samples were sequenced as paired-end 42 bp reads, whilst ChIP-seq samples were sequenced as single-end 75 bp reads.

### RNA-seq data processing and differential gene expression analysis

Standard data analysis was carried out by Active Motif. Briefly, for HDLEC, fastq files were aligned to the hg38 reference human genome using STAR v2.4.2a^49^, with default parameters. Reads were retained if both paired-end reads aligned to the same chromosome and same strand. Gene counts were quantified using featureCounts from the subread package (v1.5.2)^50^, using NCBI RefSeq gene annotations adapted by merging overlapping exons from the same gene to form a set of disjointed exons for each gene and by merging genes with the same Entrez gene identifiers. Differential gene expression was calculated using DESeq2 v1.14.1 with a Wald test to determine significance^51^ and differentially expressed genes were defined as having an adjusted p-value < 0.05 and absolute log_2_ fold-change > 0.5. For HUVEC, fastq files were aligned to hg19 using STAR v2.4.2a. FeatureCounts from the subread package was used with the minOverlap set to 25. The number of fragments overlapping predefined genomic features of interest, e.g. genes, were counted. Only read pairs that had both ends aligned were counted. Read pairs that have their two ends mapping to different chromosomes or mapping to the same chromosome but on different strands were discarded. Overlap of differentially expressed genes between the two cell types was calculated based on overlap of unique NCBI, formerly Entrez, gene IDs.

### ATAC-seq and ChIP-seq data processing

For ATAC-seq and ChIP-seq data, FastQC v0.12.1 ^52^ was used to assess raw data quality and reads were trimmed using fastp v0.12.4 ^53^. Fastp was run with the --detected_adapter_for_pe and -l 20 parameters for paired-end ATAC-seq data and with the -l 10 parameter for single-end ChIP-seq data. Trimmed reads were aligned to GRCh38 (GCA_000001405.15_GRCh38_no_alt_analysis_set) using bowtie2 v2.4.4 ^54^ with the parameters --very-sensitive (ChIP-seq) or --very-sensitive --no-mixed --no-discordant -I 20 -X 700 (ATAC-seq). Post-alignment QC reports were generated using qualimap v2.2.2 ^55^. Aligned files were processed with samtools v1.14 ^56^ to remove reads which were duplicates, unmapped, which failed the platform quality checks or had an alignment quality < 30. Mitochondrial reads were removed from ATAC-seq data. ENCODE blacklisted regions obtained from https://github.com/Boyle-Lab/Blacklist^57^ were removed from both datasets using bedtools v2.30.0 intersect^58^. Alignment statistics are provided in **Supplementary Table 1**.

For HDLEC ATAC-seq (n = 2), alignment coordinates were shifted to correct for transposase insertion using deepTools v3.5.1 alignmentSieve^59^ with the --ATACshift flag. We first called peaks in individual replicates using macs2^60^ with the following parameters: -f BAMPE -- nomodel --shift --broad --shift 37 --extsize 73 --keep-dup all --cutoff-analysis -p 1e-3. Individual replicate peaks (macs2 q < 0.05) were then used to define *‘HDLEC accessible regions’* using Irreproducible Discovery Rate (IDR)^61^. The IDR cut-off was set to 0.05. For HUVEC ATAC-seq, replicated peaks for the larger number of replicates (n = 5) were defined as those found in a pooled peak set (macs2 with Benjamini-Hochberg corrected p-value, or q-value < 0.01) and at least two individual replicates (macs2 with q < 0.05), which returned a similar number of replicated peaks to those defined in HDLEC using the IDR method. For HDLEC ChIP-seq data, ERG and H3K27ac peaks (both n = 1) were called using macs2 with the parameters -B --keep-dup all -f BAM, with a stringent threshold q < 0.001 (**Supplementary Table 5**). Note that q < 0.001 was chosen following the comparison of several q-value thresholds to obtain a “high-confidence” list of peaks for initial study of the ERG binding landscape and target genes in HDLEC.

### Classification of HDLEC cis-regulatory elements (CRE)

HDLEC CREs, defined as accessible chromatin regions (IDR ATAC-seq peaks, see previous section) were classified as ‘promoters’, ‘enhancers’ and ‘other’ as described: CREs intersecting H3K27ac peaks were defined as promoters or enhancers based on the presence or absence of a HDLEC CAGE peak respectively^39,62^; the remaining CREs were labelled as ‘other’ (CREs coordinates are provided in **Supplementary Table 2**). CAGE peaks were retrieved from the FANTOM6 data download site (https://fantom.gsc.riken.jp/6/datafiles/) and those corresponding to HDLEC (n = 3) were extracted and filtered for peaks with count > 0 in all three donors (n = 36,409 in total). Heatmaps showing ATAC-seq, H3K27ac ChIP-seq and CAGE signal were generated using deepTools computeMatrix and plotHeatmap with pooled coverage bigwigs as input. The reference-point was set as the centre coordinate of the ATAC-seq peaks, with bin sizes of 10 bp and 2 kb extensions up- and down-stream. The ChIPseeker package v1.28.3 ^63^ was used to annotate the genomic distribution of open chromatin regions and ERG binding sites at promoters, 5’-UTRs, exons, introns and intergenic regions. ChIPseeker was used with pre-computed annotations from the TxDb.Hsapiens.UCSC.hg38.knownGene Bioconductor package.

HDLEC super-enhancers were defined using the ROSE algorithm v0.1 ^64,65^, with HDLEC enhancers and H3K27ac ChIP-seq coverage as input. Default parameters and a transcription start site exclusion zone size of 5 kb (“-t 2500”) was used for ranking the stitched regions. Putative target genes were assigned using Genomic Regions Enrichment of Annotations Tool (GREAT)^66^ with default parameters (basal plus extension: proximal distance 5 kb upstream and 1 kb downstream, plus distal up to 1000 kb).

### Transcription factor motif enrichment analysis

The enrichment of transcription factor motifs at HDLEC CREs was calculated using Hypergeometric Optimization of Motif EnRichment (HOMER) v4.11 ^67^ with parameters: -size given -mask -bits -cpg and the whole genome (hg38) set as the background.

### EC subtype-specificity analysis

ERG binding sites were classified into those specific to HDLEC, specific to HUVEC, and shared between the two cell types. EC subtype-specific ERG binding sites were defined using individual replicate “high-confidence” peaks (i.e., those called using macs2 with a strict significant threshold, q < 0.001) which were absent from the list of “permissive” peaks in the alternative cell type (q < 0.05). Shared peaks were defined as any peaks called in HDLEC or HUVEC with q < 0.001 which were not in the list of HDLEC-specific or HUVEC-specific peaks, defined above. HDLEC and HUVEC open chromatin regions were classified as cell type-specific or shared based on the intersection of LEC IDR ATAC-seq peaks and replicated HUVEC peaks (see section *ATAC-seq and ChIP-seq data processing*). Shared sites were calculated as the number of HDLEC peaks which intersected with a peak in HUVEC. Heatmaps were generated using *deepTools computeMatrix* for individual samples, with the final ATAC-seq heatmap plotted using the median signal per 10 bp bin across replicates.

### Functional annotation analyses

Putative target genes with ERG binding sites were assigned using rGREAT v2.2.0 ^68^ with default parameters (basal plus extension: proximal distance 5 kb upstream and 1 kb downstream, plus distal up to 1000 kb). These genes were filtered for differential expression (FDR < 0.05 and absolute log2FC > 0.5) following siRNA ERG knockdown in HDLEC to prioritise genes likely regulated in *cis* by ERG. The lists of putative target genes were input to EnrichR^69–71^, a gene set search engine to investigate the enrichment of biological process gene ontology (GO) terms.

### Transcription factor co-occurrence

To explore TF co-occurrence, raw ChIP-seq data for PROX1, GATA2, FOXC2 and NFATC1 in HDLEC^42^ were downloaded using the fasterq-dump command from sra-tools v2.11.0. The ChIP-seq data was then processed as described above and peaks were called together with ERG using a more lenient macs2 threshold of q < 0.05 to carry out a more sensitive comparison of the global overlap of binding profiles. Coverage bigwig files were generated using deeptools bamCoverage with the parameters --binSize 10 --normaliseUsing -- extendReads 200 --ignoreForNormalization chrX.

Co-occurrence of ERG, PROX1, GATA2, NFATC1 and FOXC2 was first explored at HDLEC CREs by intersecting CREs with ChIP-seq peaks using *bedtools intersect*. The list of CRE intersections was input to TF-COMB (Transcription Factor Co-Occurrence using Market Basket analysis; v1.0.3)^43^ which calculates cosine scores of association and Z-scores of significance by shuffling TF names rather than coordinates to account for the natural occurrence of TFs in clusters^43,72^. TF-COMB parameters were set as max_dist = 200 and max_overlap = 1. Genome-wide TF overlap was also directly quantified from ChIP-seq peaks using bedtools intersect and the *upset* command from the *UpSetR* package^73^. We also investigated the effect of additional TFs binding together with ERG on the cell type specificity of target gene expression. HDLEC CREs bound by ERG, or ERG+1, ERG+2, ERG+3 or ERG+4 TFs were defined using *bedtools intersect* and putative target genes were assigned using rGREAT, subset for genes with a median FPKM >1 across three replicates. An independent series of RNA-seq datasets were used to quantify gene expression in HDLEC compared with other endothelial cell types (**Supplementary Table 1**) including HDLEC, HUVEC, liver sinusoidal ECs (LSEC), aortic and pulmonary artery ECs, lymphatic-specific and blood-specific endothelial-colony forming cells (ECFCs) and corneal endothelial cells. Briefly, raw data were downloaded and processed in-house with a consistent pipeline including read trimming (fastp v0.12.4), alignment to the GRCh38 reference genome (STAR v2.7.9a) and quantification (salmon v1.5.2 with the --gcBias parameter) based on the GENCODE v36 reference transcriptome^74^. HDLEC-enriched expression was calculated as Z-scores and plotted in R using the Python Seaborn style via the reticulate package v1.35.0.

### Data visualisation

Visualisation of RNA-seq and epigenomic datasets at candidate loci was carried out using the UCSC Genome Browser (GRCh38)^75^. Custom tracks show coverage normalised by Bins Per Million mapped reads (BPM). Pooled coverage is shown for assays with multiple replicates, where each represents a different donor.

### Quantitative real-time polymerase chain reaction

Total RNA was extracted from cultured cells using the RNeasy Kit (Qiagen). Complementary DNA (cDNA) was synthesised using Superscript III Reverse Transcriptase (Thermo Fisher Scientific). Quantitative real-time PCR (qRT-PCR) was carried out using PerfeCTa SYBR Green FastMix (Quanta Bioscience) on a Bio-Rad CFX96 thermocycler. Relative quantification of gene expression was carried out using the comparative cycle threshold method (ΔΔCt) with *GAPDH* as the reference gene. See **Supplementary Table 10** for list of oligonucleotides used in this study.

### Immunoblotting

Cell lysates were collected using RIPA buffer (Sigma-Aldrich) containing 1% protease and phosphatase inhibitor cocktail (Sigma-Aldrich). Whole protein lysates were separated using Bolt Bis-Tris Plus Mini Protein Gels (4-12%) and transferred to polyvinylidene difluoride membrane (PVDF) using iBlot 2 western transfer system (Thermo Fisher Scientific). Primary and secondary antibodies used are listed in **Supplementary Table 11**. Detection and quantification of fluorescence intensity was carried out using an Odyssey CLx Imaging System (LI-COR Biosciences, Cambridge) and Image Studio software (Ver.4; LI-COR).

### Proximity-Ligation Assay (PLA)

HDLEC were seeded onto 8-well Nunc Lab-Tek II Chambered Coverglass (Thermo Fisher Scientific) and transfected with 20 nM control or ERG siRNAs for 48 hr. HDLEC were fixed with 4% paraformaldehyde for 12 mins, washed with PBS and then permeabilised for 15 mins using 0.5% Triton X-100 in PBS. Duolink Proximity Ligation Assay (PLA; Sigma-Aldrich) was performed according to the manufacturer’s instructions. The following primary antibodies were used for the PLA: mouse or rabbit anti-ERG, goat anti-PROX1 and mouse anti-GATA2. Secondary antibodies conjugated with oligonucleotides were anti-goat or anti-rabbit PLUS probes and anti-mouse MINUS probe (Sigma-Aldrich). Relevant host species IgG were used as negative controls. PLA signal was detected using Duolink In Situ Detection Reagent Red (DUO92008; Sigma-Aldrich). After the last wash using Duolink Wash Buffers (DUO82049; Sigma-Aldrich), slides were mounted using Fluoroshield Mounting Media with DAPI (ab104139; Abcam, Cambridge). PLA signals were imaged using confocal microscopes LSM780 (Carl Zeiss) and SP8 (Leica).

### Matrigel in vitro tubulogenesis assay

Cells (1 × 10^4^) were seeded onto µ-slide Angiogenesis (Ibidi, Gräfelfing, Germany) precoated with Matrigel-Reduced Growth Factor Matrix (Corning). Tube formation was examined after 24 hr using a 10X objective lens on an EVOS XL Core microscope (Thermo Fisher Scientific). Images were analysed using WimTube Image Analysis Software (ibidi).

### HDLEC spheroid 3D sprouting assay

EC spheroids were generated using a hanging drop method^76^. Briefly, cells (4 × 10^4^) were suspended in complete media containing 0.24% (w/v) methyl cellulose 4000 cP (Sigma-Aldrich). Drops of 20 µL cell suspension were incubated upside-down for 24 hr at 37 °C in 5% CO_2_ to allow spheroid formation. EC spheroids were collected by centrifugation and embedded in 6% (w/v) gelatin hydrogels modified with a 1:2 ratio mix of tetrazine and norbornene, as previously described^77^. Gelatin-embedded EC spheroids were incubated for 24 hr in complete media. In some experiments, HDLECs were incubated in serum-reduced media (basal media supplemented with 0.1% FBS, 0.2 µg/mL hydrocortisone and 1 µg/mL ascorbic acid). ECs in serum-reduced media were starved for 4 hr before the addition of VEGF-C (50 ng/mL; R&D Systems). After 24 hr, brightfield images were obtained using an inverted microscope DMi1 (Leica Microsystems) at 10X magnification, to quantify the spheroid core area, sprouts length and number using ImageJ (NIH). Cells were then fixed in 4% (w/v) paraformaldehyde (Thermo Fisher Scientific) and permeabilized using 0.1% (v/v) Triton X-100. Cells were stained using the nuclear counterstain Hoechst 33342 (Thermo Fisher Scientific) and rhodamine phalloidin (Thermo Fisher Scientific) to identify actin filaments. Z-stack confocal images were obtained using a Leica SP8 confocal microscope.

### Proliferation assay

Cell proliferation was assessed using Click-iT Edu Proliferation Assay (C10499; Thermo Fisher Scientific), according to the manufacturer’s instructions. Briefly, 1 ×10^4^ cells per well were seeded onto a 96-well plate in complete media. HDLEC were pre-treated with 20 nM control or ERG siRNAs for 24 hr. Cells were incubated with 10 µM Edu (5-ethynyl-2’-deoxyuridine) for 4 hr in a humified incubator at 37 °C in 5% CO_2_. The fluorescent signal generated by the Amplex Ultra Red reagent was detected with excitation 568 nm and emission 585 nm using a CLARIOStar Plus plate reader (BMG Labtech). Values of each sample were normalised to background signal from control wells without Edu treatment.

### Animal experiments

Generation of *Erg^flox/flox^* mice has been reported previously^20^. *Erg* floxed mice were crossed with a transgenic mouse line expressing tamoxifen inducible Cre recombinase (*CreER^T^*^2^) under the control of the vascular endothelial cadherin (*Cdh5(PAC)*) promoter^78^ or the *Prox1* gene promoter, kindly provided by Tajia Makinen^32^. These lines were maintained on a C57BL/6J background. Control mice were *ERG^flox/flox^*littermates that were negative for *CreER^T^*^2^. Cre-mediated recombination was induced by intraperitoneal injection of 50 µg of tamoxifen (T5648; Sigma-Aldrich) dissolved in peanut oil (1 mg/ml) at postnatal days (P)0–P3. Mice were analysed 21 days after tamoxifen administration. Both male and female mice were used for experiments. All animal experiments were conducted with ethical approval from Imperial College London in compliance with the UK Animals (Scientific Procedures) Act of 1986. Mice were housed in a pathogen free facility and kept in a 12-hr light/dark cycle in a controlled temperature room (20.5-23.5 °C and 50-60% humidity).

### Immunofluorescence analysis of cells and mouse tissues

Cells were fixed with 4% (w/v) paraformaldehyde (Sigma-Aldrich) for 15 min, then permeabilised for 5 min with 0.5% Triton X-100 (Sigma-Aldrich) before blocking with 3% (w/v) bovine serum albumin (BSA; Sigma-Aldrich) for 1 hr. Cells were incubated with primary antibodies diluted in blocking buffer overnight at 4 °C. Secondary antibodies conjugated to Alexa Fluor dyes (Thermo Fisher Scientific) were diluted 1:500 in blocking buffer and incubated with cells for 1 hr at room temperature. Primary and secondary antibodies are listed in **Supplementary Table 11**. Nuclei were visualised with 4ʹ,6-diamidino-2-phenylindole (DAPI; Thermo Fisher Scientific). Confocal imaging was carried out using Zeiss LSM780 confocal microscope and Zen software or a Nikon A1R point scanning confocal microscope. Images were analysed with ImageJ (NIH) or Volocity (Quorum Technologies).

Whole-mount immunofluorescence of mouse skin was carried out as previously described^47^. Briefly, mouse ears were separated by forceps and the dorsal side was processed for staining. Samples were fixed in 4% paraformaldehyde, permeabilised in 0.3% Triton X-100 in PBS (PBST) and blocked with 3% milk in PBST. Primary antibodies were diluted in blocking buffer and incubated with the samples overnight at 4 °C. Samples were washed several times using PBST and incubated for 1 hr at room temperature with fluorescence-conjugated secondary antibodies in blocking buffer. Samples were washed several times with PBST and mounted onto glass slides using Fluoromount-G (Thermo Fisher Scientific). Images were acquired using Zeiss LSM 780 confocal microscope and Zen software. Images of tissues and cells represent maximum intensity projections of Z-stacks.

### ERG variants identification

Individuals diagnosed with primary lymphoedema, who had tested negative for the R136 primary lymphoedema gene panel (https://panelapp.genomicsengland.co.uk/panels/65/), were enrolled in the lymphatic disorder subdomain in the Cardiovascular Genomics England Clinical Interpretation Partnership (GeCIP) of the Genomics England (GEL) 100,000 Genomes Project Rare Diseases program. In addition to the 4 high impact variants we previously identified^16^, we sought to identify novel cases with missense variants. For the analysis of the whole genome sequencing data the main program_v11 in GEL was used to identify genomes with “Primary Lymphoedema” label (but lacking confirmed causal variants in known PL genes). Genomics England aggV2 files were interrogated to identify participants with rare (gnomad MAX MAF<0.0001) missense variants in *ERG*, predicted damaging by the Combined Annotation Dependent Depletion (CADD) tool^79^ (CADD PHRED-like score >15). Missenseimpacting on the canonical transcript were considered.

### Variant in silico analysis

The relative genomic and protein positions of ERG reported in this study correspond to ERG transcript 1 (also known as ERG-3, P55), annotated in NCBI GenBank under accession RefSeq: NM_182918.4; Ensembl: ENST00000288319.7; UniProt: P11308-4. The reported genomic coordinates refer to the GRCh38/hg38 human genome reference. Putative changes in the ERG gene structure and/or amino acid sequence caused by the reported variants were retrieved from the University of California–Santa Cruz (UCSC) refGene database. Allele frequencies (AF) were checked in gnomAD v4 ^80^. Pathogenicity was predicted using Combined Annotation Dependent Depletion (CADD)^79^, Rare Exome Variant Ensemble Learner (REVEL)^81^, and spliceAI^82^ scores. The six rare variants identified in probands with primary lymphoedema overlapping exonic regions of *ERG* were further investigated using MetaDome^27^. In brief, MetaDome genetic tolerance profiles were previously derived using 56,319 transcripts, 71,419 protein domains, 12,164,292 genetic variants from gnomAD^25^ and 34,076 pathogenic mutations from ClinVar^83^ and queried via the online portal at https://stuart.radboudumc.nl/metadome.

Where DNA was available, Sanger sequencing validation and co-segregation analysis were carried out. Primers were designed using Primer3 program^84^. PCR products were sequenced using Source Bioscience (Cambridge) services. The sequencing traces were analysed using Finch TV v1.4 (Geospiza, Inc, Seattle, WA, USA) and CLC Sequence Viewer 6.4 (CLC bio A/S).

### Participants

This study has Research Ethics Committee approval (REC ref: 14/LO/0753) and informed consent was obtained for all participants. Written consent for publication of patient images has been obtained and archived in our site files.

Blood samples (48 ml) were collected from healthy volunteers (not diagnosed with primary lymphoedema) and a patient with primary lymphatic anomaly. Peripheral blood mononuclear cells were isolated from blood samples collected in Vacutainer CPT tubes (BD Biosciences, Wokingham) containing sodium heparin anticoagulant. Cells were seeded at a density of 3-5 × 10^7^ cells per well in complete endothelial growth medium (EGM)-2 (Lonza, Slough), supplemented with 20% (v/v) Human Serum from Male AB plasma (Sigma-Aldrich) onto six-well plates pre-coated with type I rat tail collagen (Corning, Amsterdam) and incubated in a humidified chamber at 37 °C in 5% CO_2_, as previously described^85^. After 24 hr, nonadherent cells and debris were aspirated, adherent cells were washed once with EGM-2 medium, and fresh EGM-2 was added to each well. Medium was changed daily for 7 days and then every 2 days. Discrete ECFC cobblestone monolayer colonies appeared between 10-15 days in culture. ECFC derived from the colonies were sub-cultured and characterised for endothelial morphology. Experiments were performed using ECFC between passages 4 to 6.

### Molecular Dynamics Simulations

The molecular dynamics (MD) models relied on the available crystal structure of the ERG ETS Domain-DNA complex, which has been solved to 2.77 Å resolution using X-ray diffraction (PDB-ID: 4iri)^28^. This structure was used as a starting point to model wild-type ERG in the presence or absence of DNA. The DNA sequence used in the MD model is the same sequence as in the crystal structure PDB 4iri^28^. To model the ERG variant p.Y388C, the residue sequence at position 388 was changed using UCSF Chimera^86^. Using Assisted Model Building with Energy Refinement (AMBER) software package with the ff14SB force field^87,88^, the structures were solvated in a rectangular box of TIP3P water molecules with a minimum distance of solute molecules to the box surface of 10.0 Å and 12.0 Å for the simulations in the absence and presence of DNA, respectively. Five repeat simulations were run for each model (ERG wild type and p.Y388C variant, with and without DNA) for 3 μs duration. The simulations in explicit conditions followed the following series of steps: 1) Energy minimisation: an initial 1000 cycle minimisation of the solvent was carried out switching from steepest descent to the conjugate gradient algorithm after 100 cycles. The solute molecules were restrained using harmonic restraint with a force-constant of 1 kcal mol^−1^ Å^−2^. Next, a minimisation for the entire system was performed for 5000 cycles, switching minimisation algorithms after 500 cycles. 2) Heating: the system was heated over 62.5 ps to 100 K using a linear temperature gradient in the constant volume ensemble (NVE ensemble). Next, the system was heated to 310 K over 300 ps with a linear temperature gradient in the constant pressure ensemble (NPT ensemble), followed by a 200 ns equilibration with isotropic pressure scaling. 3) Equilibration: structure equilibrations were carried out at 310 K using constant pressure and temperature (NPT ensemble) with isotropic pressure scaling using a 2.5 fs time-step. The SHAKE algorithm was used to restrain hydrogen bonds. Particle mesh Ewald (PME) boundary conditions were used to calculate long-range electrostatics with 8 Å cut-off for non-bonded interactions. Temperature was controlled using Langevin dynamics with a collision frequency set at 1 ps^−1^. To maintain the pressure at 1 bar a Monte Carlo barostat was used with volume changes occurring every 100 steps. The trajectory was analysed over the last 1.5 μs with a step size of 10 frames. Data were produced with UCSF Chimera and Python, including the Matplotlib and Seaborn packages^89,90^. For the boxplots, the box represents the standard deviation between the runs and the line in the middle shows the average value of all runs.

### Plasmids and site-directed mutagenesis

Human ERG cDNA (NCBI Accession NM_182918), modified to express a C-terminal 6X His epitope tag, was cloned into the mammalian expression vector pcDNA3.1 (Invitrogen). A 1.078 kb region of the *THBD* promoter proximal to the transcription start site cloned into the pGL4.10[luc2] luciferase reporter vector (Promega, Madison, USA) was previously described^23^. Firefly luciferase empty vector pGL4.10[luc2] was used as a control; *Renilla* luciferase vector pGL4.74 [hRluc/TK] vector] (Promega) was used as an internal control in the dual-reporter assay.

Mutation of *ERG* cDNA to replicate the *ERG* coding variants found in patients with PL was carried out using QuickChange Lightning Site-directed Mutagenesis Kit (Agilent, Stockport), according to the manufacturer’s instructions. Sequences of the mutagenic oligonucleotides are listed in **Supplementary Table 10.** Cloning was confirmed by Sanger sequencing (Azenta Life Sciences, Abingdon). For *ERG* variant p.N463T, the ERG cDNA sequence was synthesised using GeneArt (Thermo Fisher Scientific), and then cloned into pcDNA3.1.

### Reporter assays

HEK293 cells (2 × 10^4^ cells), seeded onto 6-well plates, were co-transfected with cDNA to wild type ERG or *ERG* variants in pcDNA3.1 expression plasmids along with luciferase reporter plasmids (1 µg each) using 150 mM NaCl solution containing polyethyleneimine (PEI) (Polyscience, #23966) at 2.5:1 ratio to DNA. Luciferase activity was measured after 24 hr using the Dual-Luciferase Reporter Assay System (E1910; Promega) and a CLARIOStar Plus plate reader (BMG Labtech). Luciferase reporter activity was normalized to the internal *Renilla* luciferase control and is expressed relative to empty vector control.

### Electrophoretic Mobility Assays (EMSA)

HEK293 cells were transfected with cDNA of wild-type *ERG* or *ERG* variants cloned into pcDNA3.1. After 24 hr, whole cell lysate was collected in RIPA buffer containing 1% protease inhibitor cocktail (Sigma-Aldrich). EMSA was carried out using the LightShift™ Chemiluminescent EMSA Kit (20148; Thermo Fisher Scientific) according to the manufacturer’s instructions. Briefly, binding reactions contained 15 µg of protein lysates mixed with 20 fmol of end-labelled biotinylated oligonucleotide probe 5’-TCGACGGCCAAGCC<u>GGAA</u>GTGAGTGCC-3’, previously shown to bind ERG^29^. In some cases, an excess (4 pmol) of unlabelled oligonucleotide probe was also added to the binding reaction. For super-shift EMSA, the binding reactions were supplemented with 0.4 µg of mouse anti-ERG antibody or IgG negative control. Binding reactions were incubated at room temperature for 20 min. The reactions were then subjected to gel electrophoresis on a native 6% polyacrylamide retardation gel in 0.5X Tris-Borate-EDTA buffer (Thermo Fisher Scientific) and transferred to a positively charged nylon membrane. The biotin end-labeled DNA was detected using streptavidin-horseradish peroxidase conjugate and chemiluminescent substrate. DNA-protein interactions were imaged using ChemiDoc Imaging System (Bio-Rad).

### Statistical analysis

Unless otherwise stated, statistical analyses were performed using GraphPad Prism v9 software. Statistical tests included unpaired Student’s t-test and paired t-tests to compare two groups, and one-way ANOVA with Tukey’s multiple comparisons tests. All statistical comparisons were two-sided. Statistical analysis of sequencing datasets (RNA-seq, ChIP-seq and ATAC-seq) were carried out using appropriate software packages as defined in the Methods and figure legends. We consider a P value of 0.05 significant. Significance levels of *P < 0.05, **P < 0.01, ***P < 0.001, ****P < 0.0001 are used throughout.

### Data availability

All source data are available within the article and in the Supplementary Tables. All sequencing data generated in this study (RNA-seq, ATAC-seq, ChIP-seq) are summarised in **Supplementary Table 1** and upon publication will be publicly available at the NCBI Gene Expression Omnibus as a super-series. Software code developed for this study will be available at GitHub. Any other data are available from the corresponding authors upon reasonable request.

## Acknowledgements

Infrastructure support for this research was provided by the NIHR Imperial Biomedical Research Centre (BRC), The Wellcome Trust, Cancer Research UK and the Medical Research Council (MRC). This study received financial support from the British Heart Foundation (BHF) through grants to GMB (PG/20/16/35047, PG/17/33/32990) and AMR (RG/11/17/29256, RG/17/4/32662). Rosetrees Trust (Seedcorn2022\100269) supported GMB & DP; DP is supported by the NIHR Imperial BRC; TV is funded by BHF PhD Studentship FS/4yPhD/F/23/34202 to GMB & IC; DN is funded by a BHF PhD Studentship FS/4yPhD/F/20/34128 to IC, AMR and GMB. IC is recipient of a Sir Henry Dale Fellowship jointly funded by the Wellcome Trust and the Royal Society (224662/Z/21/Z). AS was jointly funded by EPSRC (EP/L015498/1) and BHF (RE/13/4/30184). SD, ES and PO were supported by the Swiss Federal National Fund for Scientific Research (CRSII5_177191/1). SM-A, SMa, KG, KO, SD and PO were supported by the MRC (MR/P011543/1) and BHF (RG/17/7/33217). DEA was funded by Qatar National Research Fund (QNRF) Graduate Scholarship Research Award (GSRA8-I-1-0210-21001). ADC acknowledges financial support from a UK Research and Innovation (UKRI) Future Leaders Fellowship (MR/S034757/1) and Economic and Social Research Council (ESRC) grant (ES/T013397/1). Computational analyses were performed at the Imperial College Research Computing Service (DOI: 10.14469/hpc/2232). This research was made possible through access to data in the National Genomic Research Library, which is managed by Genomics England Limited (a wholly owned company of the Department of Health and Social Care). The National Genomic Research Library holds data provided by patients and collected by the NHS as part of their care and data collected as part of their participation in research. The National Genomic Research Library is funded by the National Institute for Health Research and NHS England. We thank Prof Jorge Ferrer (Imperial College London & Centre for Genomic Regulation, Barcelona) and Dr Sílvia Bonàs-Guarch (Centre for Genomic Regulation, Barcelona) for critical reading of the manuscript.

## Author Contributions

Conceptualization: SJ, PM, SM, SMA, IC, PO, GMB Data curation: HM

Formal analysis: DP, HM, AS, KO, ES, SD, DG, JDRJ, DEA, KF, GS, SS, AM, DN, SMA, IC, PO, GMB

Funding acquisition: DP, SJ, PM, IRG, SM, AMR, IC, PO, GMB

Investigation: DP, AS, KO, ES, DEA, KF, GS, CP, SS, TV, RM, DOFG, SMA, GMB

Methodology: TM

Project administration: SMA, IC, PO, GMB

Resources: KR, JD, CT, SE, FE, VK, KG, TM, AC, IRG, SM, AMR, PO, GMB Supervision: DP, SJ, PM, KP, AC, IRG, SM, SMA, IC, GMB

Writing – original draft: DP, AS, HM, SMA, IC, PO, GMB Writing – review & editing: AMR, DP, HM, SMA, IC, PO, GMB

