## Extended data for "Loss of ERG function causes lymphatic vessel malformations and primary lymphoedema"

### Extended Data Figure 1

**a**

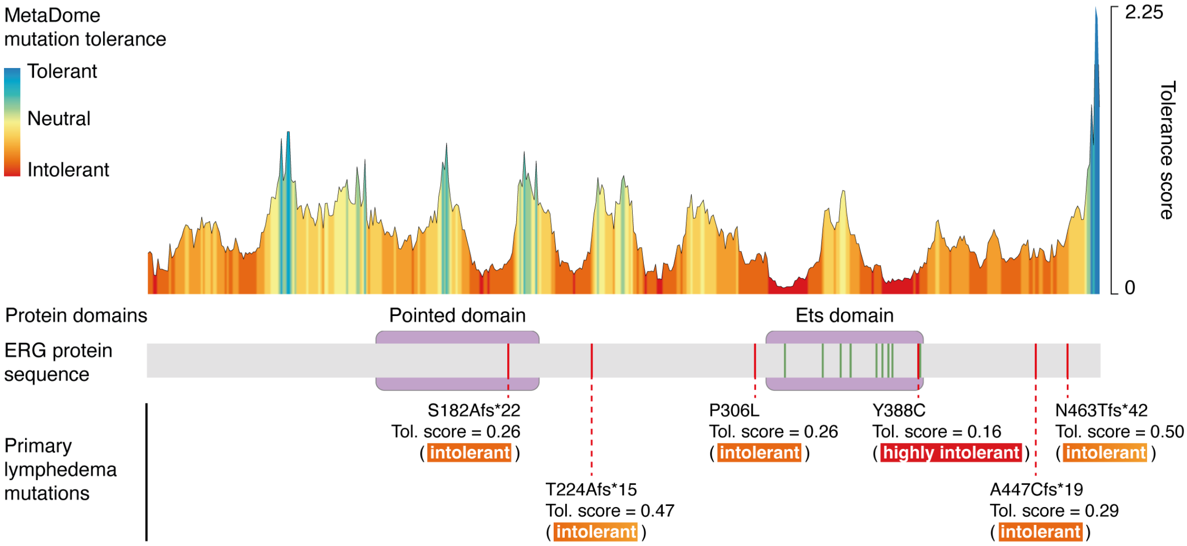


**b**

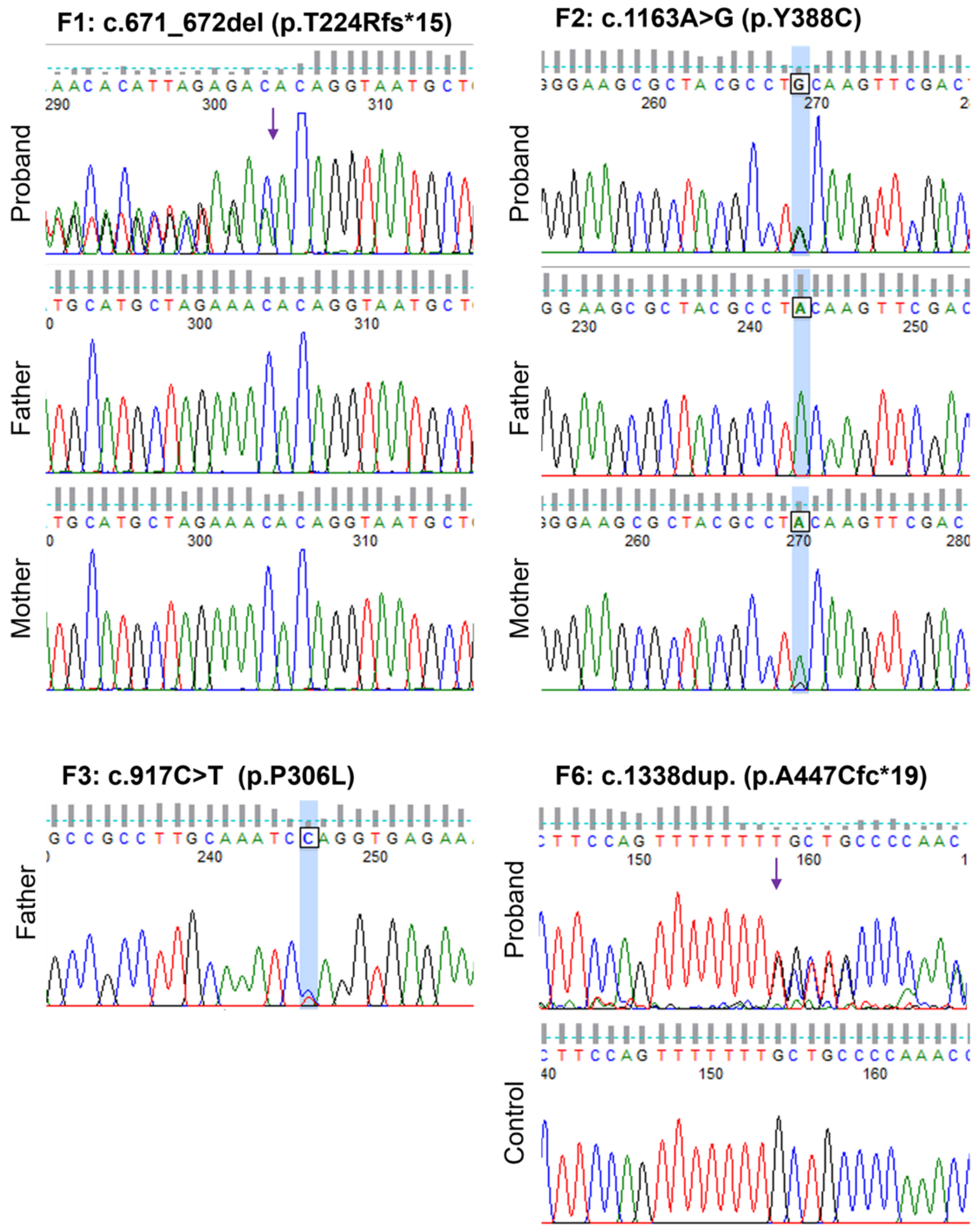

### Extended Data Figure 1

**c**

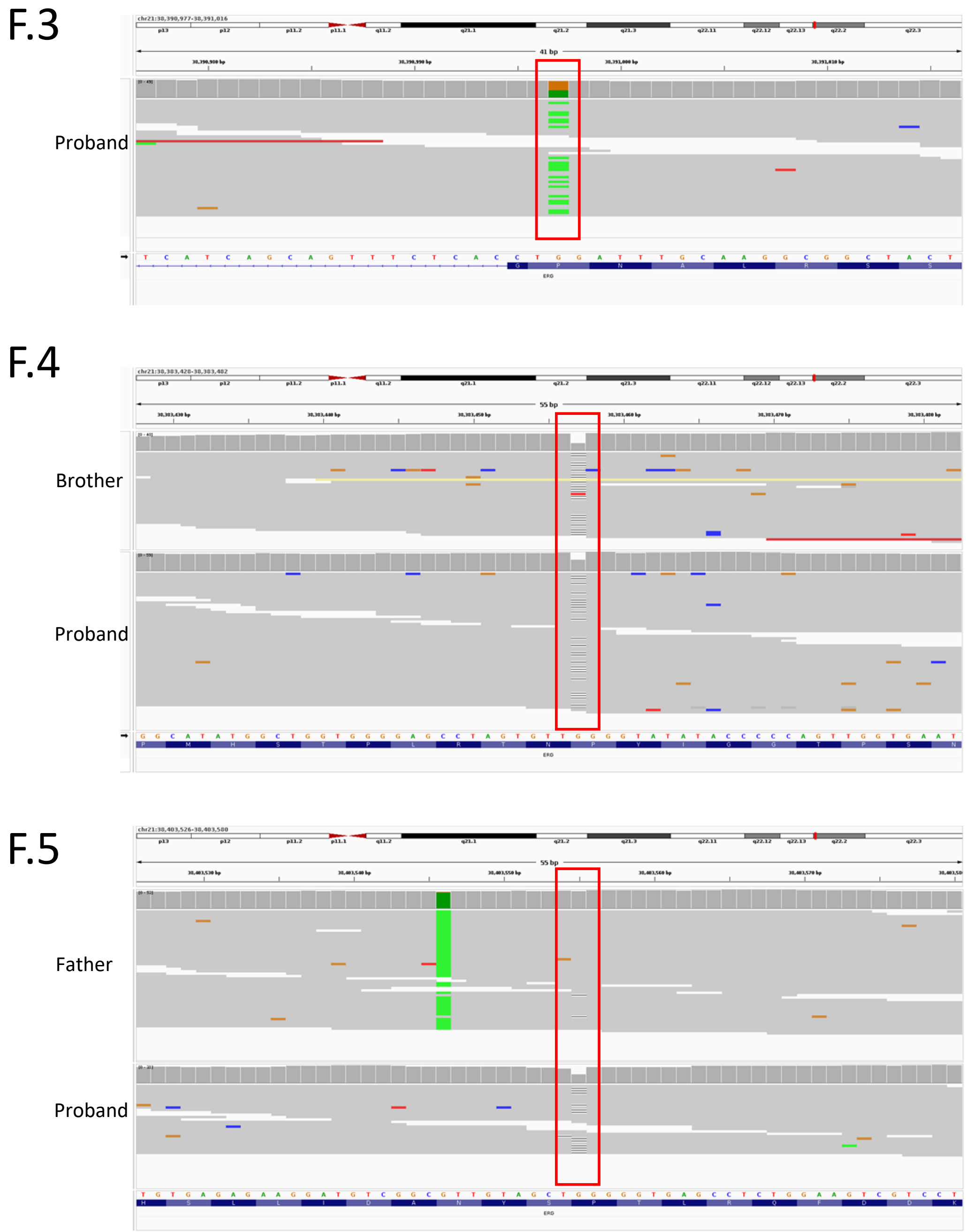


**Extended Data Figure 1. a)** Metadome analysis of six *ERG* coding variants identified in probands from the 100,000 Genomes Project. The six variants (red vertical bars) are shown relative to ERG protein meta-domains (purple boxes). Tol. score, MetaDome Tolerance Scores. **b)** Where DNA was available, Sanger sequencing and co-segregation analysis were carried out to confirm the *ERG* variants. Position of missense variants indicated with blue highlight, indels with an arrow. **c)** IGV plots of aligned sequencing data for those individuals where DNA was not available for Sanger sequencing confirmation of variants. The plots depict alignment of sequencing reads, with the variant of interest highlighted in a red box. Panel F.3: Variant (highlighted in green) at chr21:38390997 were found in 19/43 aligned reads in proband of Family 3. Panel F.4: Deletions of a single base marked with horizontal black lines at chr21:38383456 in 13/34 and 24/54 reads in the brother (top trace) and proband (bottom trace) of Family 4. Panel F.5: Deletions of a single base marked with horizontal black lines at position chr21:38403554 in 2/50 and 13/25 reads in the father (top trace) and proband (bottom trace) of Family 5.

### Extended Data Figure 2

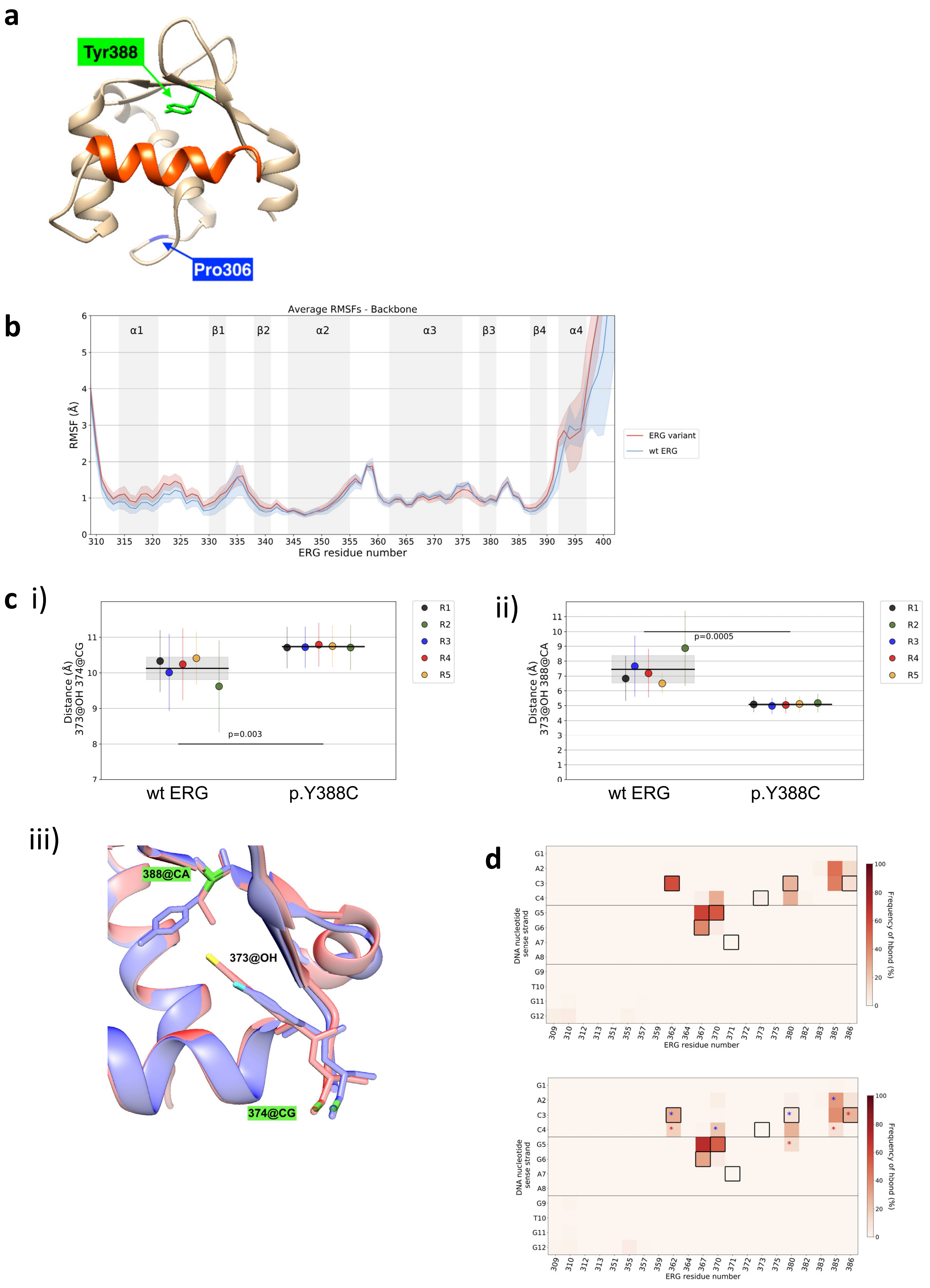

**Extended Data Figure 2. a)** Structural representations of the ETS binding domain of wild type ERG derived from the AlphaFold structure (AF-P11308), highlighting the location of two ERG variant sites, Tyr388 (green) and Pro306 (blue), with respect to the DNA binding helix (orange). **b)** RMSF comparison between ERG wild type (WT) (blue) and variant p.Y388C (red), considering only the backbone. **c)** Quantification of structural differences in the sidechain location of (i) residue Tyr373 with respect to residue Asp374 and (ii) of residue Tyr373 with respect to residue Cys388. (iii) Visual representation of atoms referred to in (i) showing the ERG variant p.Y388C structure (red) and ERG WT (blue). **d)** Frequency of hydrogen bonds established between residues of the ERG p.Y388C and DNA nucleotides in the sense strand. The boxes represent DNA-residue interactions. Asterisks represent a significant difference between the ERG WT (top) and p.Y388C variant (bottom), $p<0.05$. Blue asterisks represent a more frequent interaction in the ERG WT model, red asterisks a more frequent interaction in p.Y388C.

#### Extended Data Figure 3

**a**

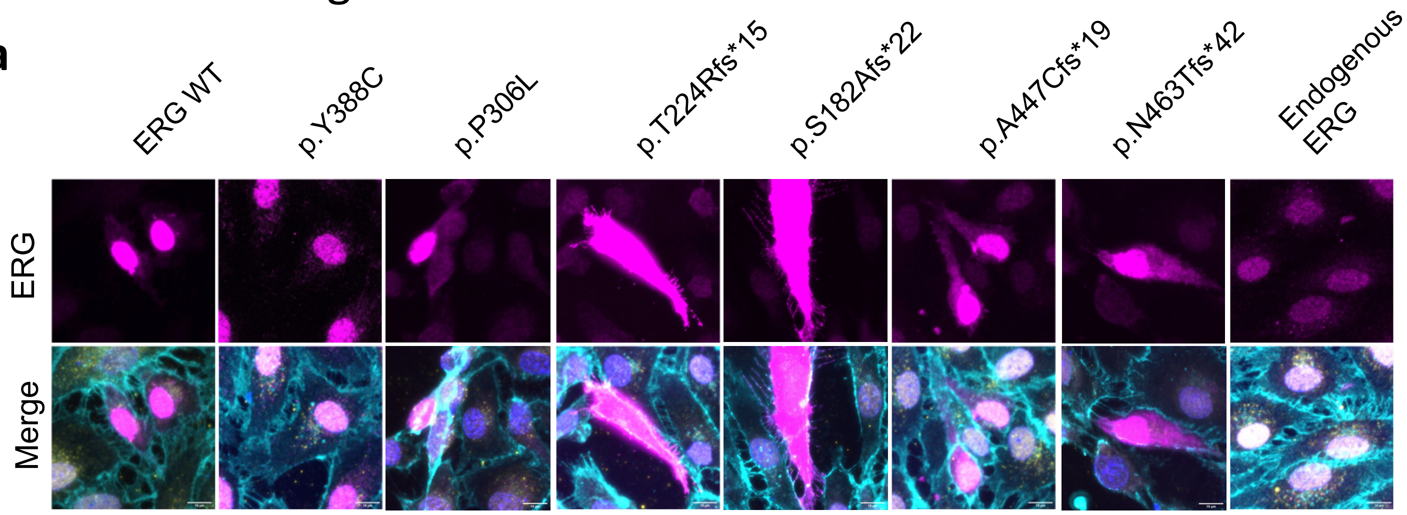


**b**

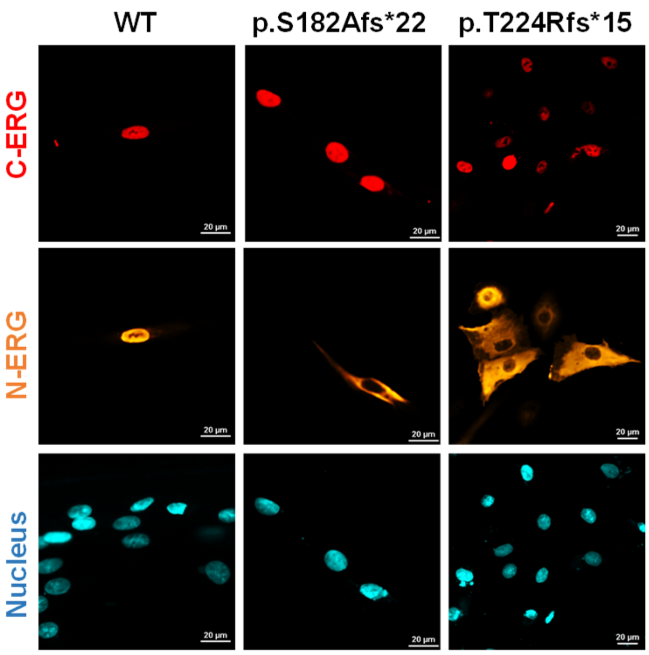


**c**

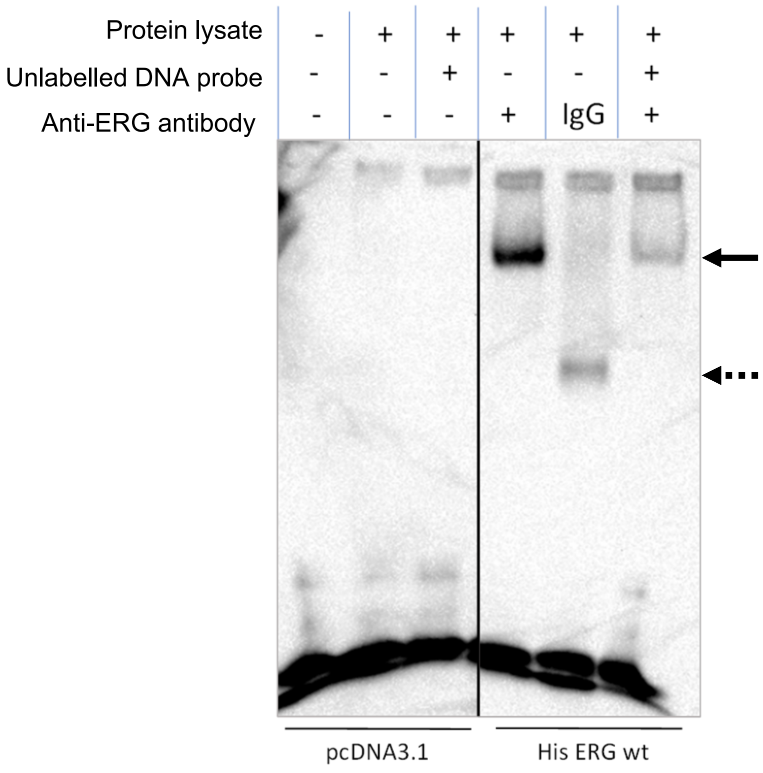

Extended Data Figure 3, continued

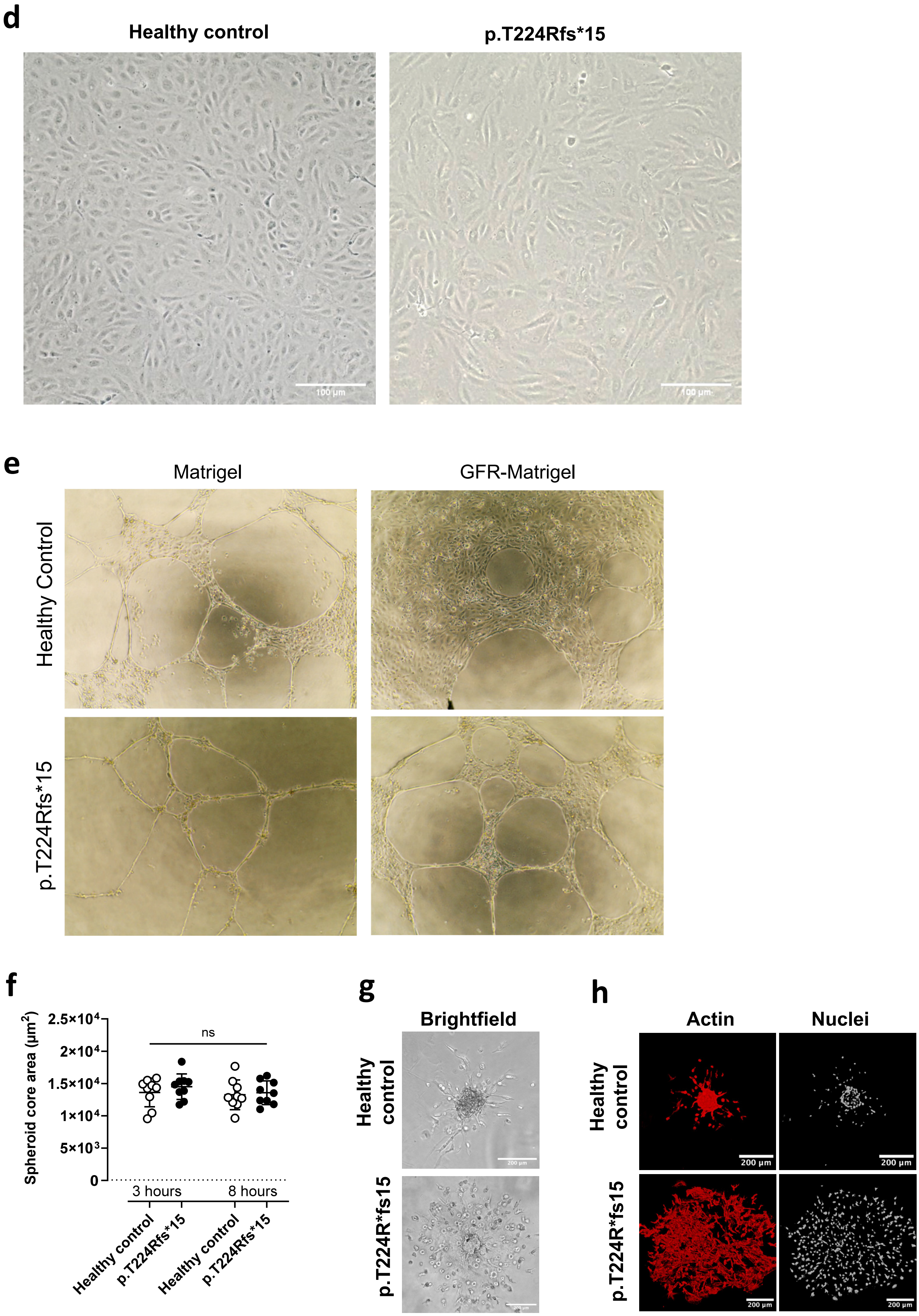

**Extended Data Figure 3. a)** Immunofluorescence microscopy of HDLEC transiently transfected with mammalian expression plasmids encoding cDNA for wild type (WT) ERG or specific ERG variants, as indicated. Cells are labelled for ERG (magenta), VE-cadherin (cyan), and nuclear staining DAPI (blue). Scale bar, 10 µm. **b)** Immunofluorescence microscopy of HDLEC transiently transfected with mammalian expression plasmids encoding cDNA for ERG wild type (WT) or specific *ERG* variants, as indicated. Cells were co-stained with antibodies recognising ERG C-terminus (C-ERG, red) or ERG N-terminus (N-ERG, orange), and nuclear marker DAPI (cyan). Scale bar, 20 µm. **c)** Representative image showing the specificity of the DNA-binding ability of WT ERG using EMSA supershift assay. ERG DNA binding was imaged using a biotinylated probe containing an ERG target sequence. The specificity of the ERG-DNA binding complex (dotted arrow) was assessed by adding anti-ERG antibody to DNA probes to induce molecular supershift (arrow), compared to IgG control or an excess of unlabelled probe. **d)** Representative light microscopy images of cultured ECFCs isolated from healthy control and a primary lymphoedema patient heterozygous for ERG p.T224Rfs*15. **e)** Light microscopy brightfield images of ECFC from healthy control and p.T224Rfs*15 seeded onto Matrigel or growth factor reduced (GFR)-Matrigel for 24 hr; 20X magnification. **f)** ECFC spheroids from healthy control and ERG p.T224Rfs*15 embedded in 6% gelatin-tetrazine/norbornene hydrogels. Quantification of spheroid sprouting assay showing spheroid core area at 3 hr and 8 hr time points; n=9 spheroids per sample/timepoint. Difference between means were analysed using a 2-way ANOVA, ns = not significant. **g)** Brightfield image of ECFC spheroids at 24 hr post embedding, scale bar 200 µm. **h)** Immunofluorescence images of ECFC spheroids at 24 hrs. Cells were labelled for actin (Phalloidin-TRITC; red) and nuclei (DAPI; greyscale). Scale bar, 200 µm.

### Extended Data Figure 4

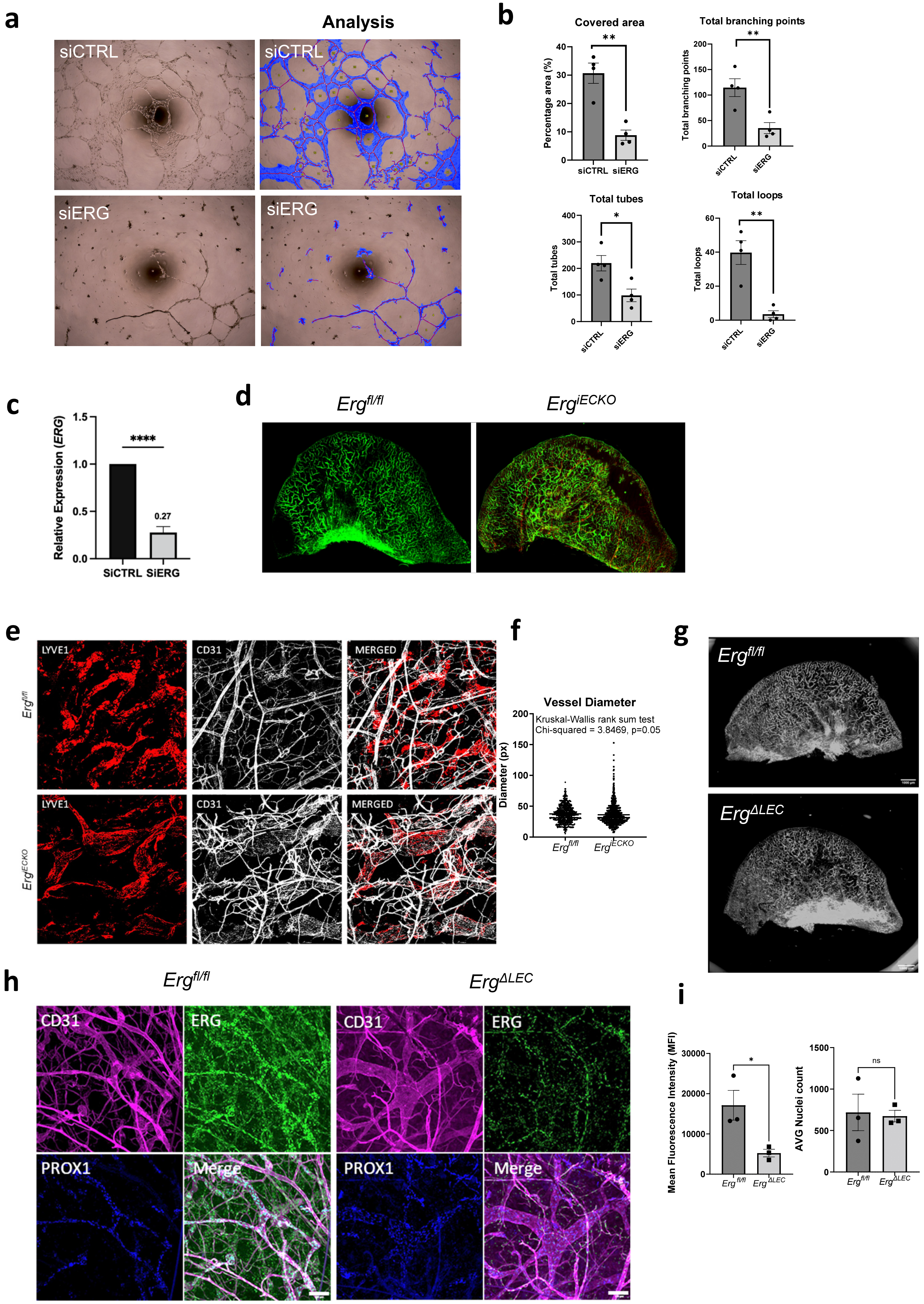

### Extended Data Figure 4, continued

**j**

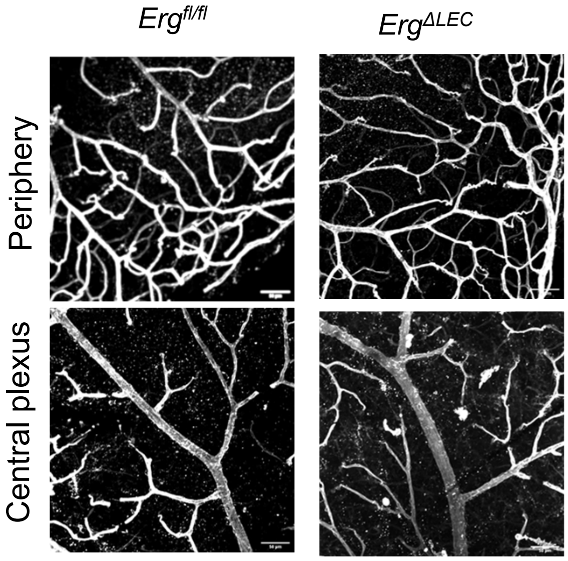


**Extended Data Figure 4. a)** Transmitted light visualisation and analysis (blue overlay) of 2D Matrigel tube formation assays using HDLEC transfected for 48 hr with non-specific control siRNA (siCtrl) and siRNA to ERG (siERG). **b)** Quantification of HDLEC tube formation: percentage area covered by tubular structures, total number of branching points, total number of tubes, and total number of loops; N=4, unpaired t-test. **c)** Relative ERG mRNA expression in HDLEC 24 hrs following siCtrl or siERG transfection was determined by RT-pPCR. Values were normalised to GAPDH and are shown relative to siCtrl-treated HDLEC, N=3, unpaired t-test. **d**) *Cdh5(PAC)CreERT2-*$Erg^{fl/fl}$ ($Erg^{iECKO}$) mice and littermate control ($Erg^{fl/fl}$) mice were treated with tamoxifen on postnatal days 1-3 and the dermal lymphatic vasculature of the ear was analysed by *en face* immunofluorescence microscopy 21 days after birth. Tissue was stained for lymphatic marker LYVE1 (green) and pan-EC marker CD31 (red). Scale bar, 1000 µm. **e**) Magnified region of ear dermis showing structures of LYVE1-stained lymphatic vessels (red) and CD31-positive blood/lymphatic vessels (grey) from $Erg^{fl/fl}$ and $Erg^{iECKO}$ mice. **f)** Quantification of lymphatic vessel diameters shows a greater range in $Erg^{iECKO}$ mice vs littermate controls, Skewness: $Erg^{fl/fl}$ = 0.39 vs $Erg^{iECKO}$ = 1.91. **g)** *Prox1-CreERT2-*$Erg^{fl/fl}$ ($Erg^{\Delta LEC}$) mice and littermate control ($Erg^{fl/fl}$) mice were treated with tamoxifen on postnatal days (P)1-3 and the dermal lymphatic vasculature of the ear was analysed by *en face* immunofluorescence microscopy P21 days after birth. Ear skin was stained for lymphatic marker LYVE1 (grey). Scale bar, 1000 µm. **h)** Ear skin from $Erg^{\Delta LEC}$ mice and $Erg^{fl/fl}$ mice was stained for CD31 (magenta), ERG (green), and PROX1 (blue). Scale bar, 100 µm. **i)** Quantification of ERG mean fluorescence intensity (MFI) and mean nuclei count per field of view; n = 3 mice, 9 fields of view per mouse; unpaired t-test. **j)** Immunofluorescent staining of mouse retina at day P21 at periphery and central vascular plexus from *Prox1-CreERT2-*$Erg^{fl/fl}$ ($Erg^{\Delta LEC}$) mice and littermate control ($Erg^{fl/fl}$) mice, labelled for IsolectinB4 (grey). Scale bar, 50 µm. All data are expressed as mean +/- SD; ns = not significant, * P < 0.05, ** P < 0.01.

### Extended Data Figure 5

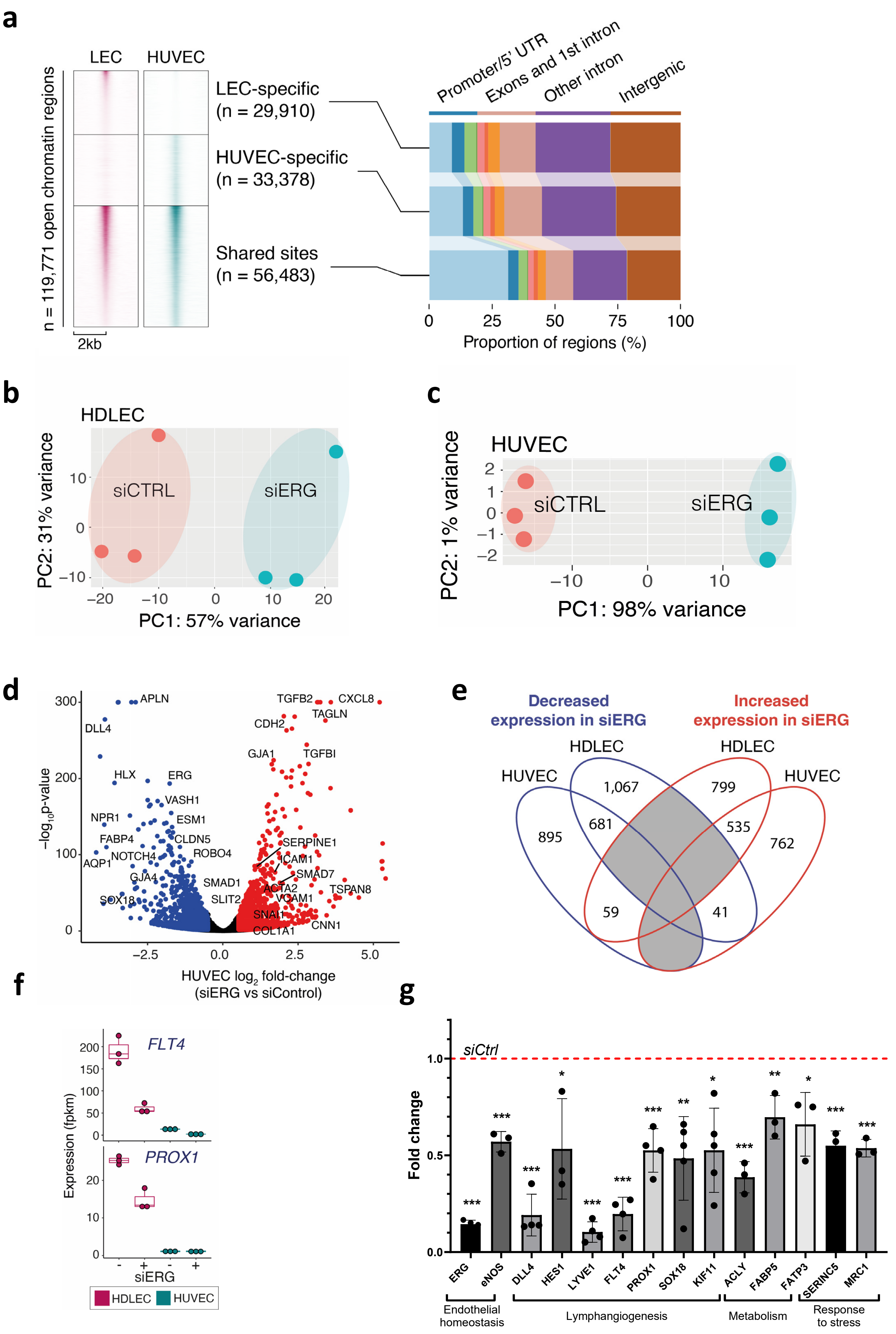

### Extended Data Figure 5, continued

**h**

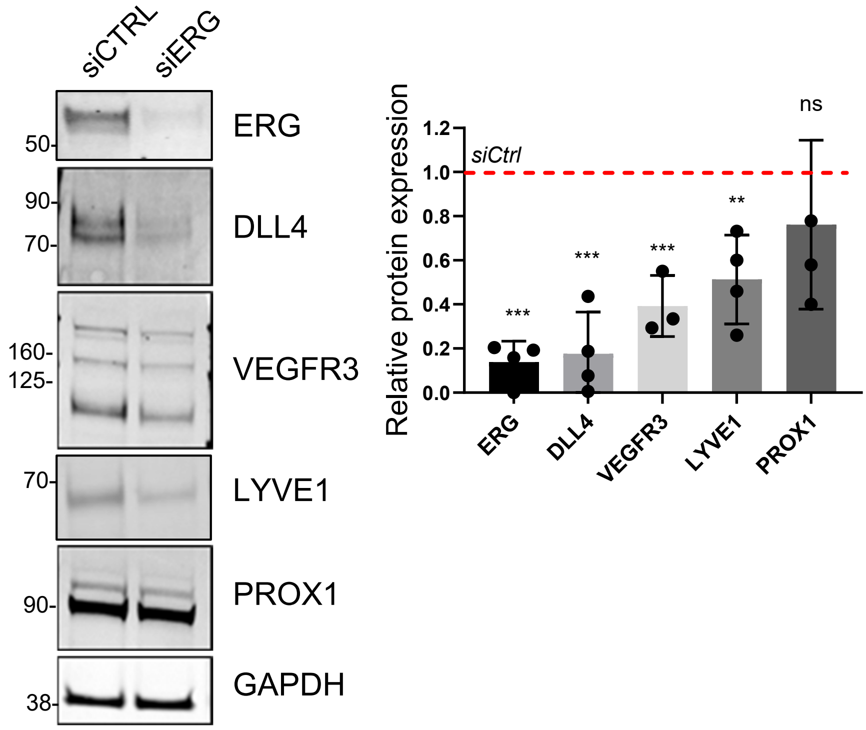


**i**

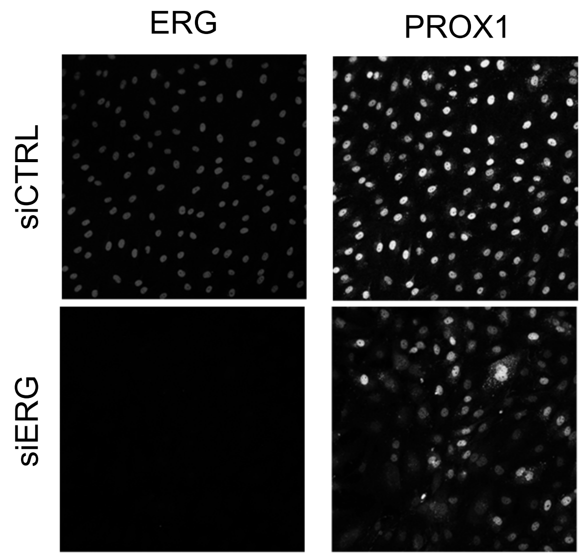


**j**

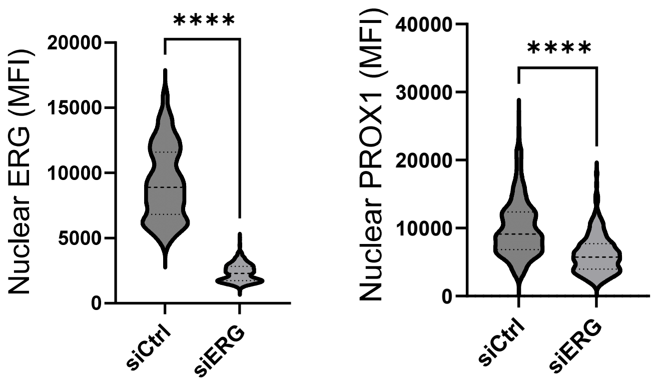


**k**

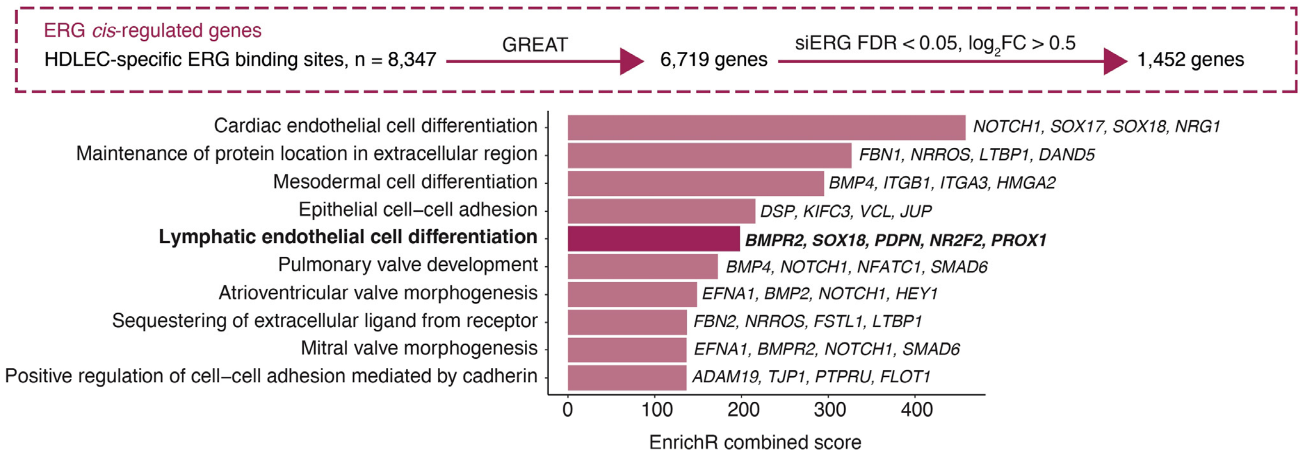

**Extended Data Figure 5. a)** (Left) ATAC-seq coverage for HDLEC and HUVEC, highlighting regions defined as replicated peaks in HDLEC-only, HUVEC-only, or both cell types. (Right) ChIPseeker functional annotation of HDLEC-specific, HUVEC-specific, and shared open chromatin regions. **b)** Principal component analysis (PCA) of RNA-seq samples in HDLEC transfected with non-specific control siRNA (siCtrl) and siRNA to ERG (siERG). **c)** PCA plot of RNA-seq samples from HUVEC treated with siCtrl and siERG. **d)** Volcano plot from RNA-seq analysis of ERG inhibition in HUVEC. Blue and red points represent genes with significantly decreased or increased expression, respectively (adjusted p-value < 0.05 and absolute log2FC > 0.5). **e)** Overlap of differentially expressed genes from siERG in HDLEC and HUVEC (adjusted p-value < 0.05 and absolute log2FC > 0.5). **f)** Boxplots showing gene expression (fpkm) of *FLT4* and *PROX1* following ERG siRNA inhibition (+ siERG) in HDLEC (magenta) and HUVEC (cyan). **g)** Message RNA expression of ERG and selected targets was determined by RT-qPCR in HDLEC transfected with ERG siRNA or non-specific control siRNA for 48 hr. Results are normalised to GAPDH and expressed as fold change relative to control siRNA (siCtrl, red line). N = 3-6, unpaired t-test. **h)** Representative immunoblots of protein lysates from siRNA treated HDLEC showing relative expression of selected proteins following ERG inhibition, normalised to GAPDH; n=3, unpaired t-test. **i)** Representative images of immunofluorescent staining for ERG and PROX1 in HDLEC transfected with siCtrl or siERG for 48h. **j)** Quantification of immunofluorescent staining assessing nuclear mean fluorescent intensity (MFI). Difference between groups were analysed using an unpaired t-test; n=3,.* P < 0.05, ** P < 0.01, *** P < 0.001. **k)** Enrichment of GO biological processes for genes annotated as ERG targets with HDLEC-specific ERG binding sites which show no evidence of ERG binding in HUVEC.

### Extended Data Figure 6

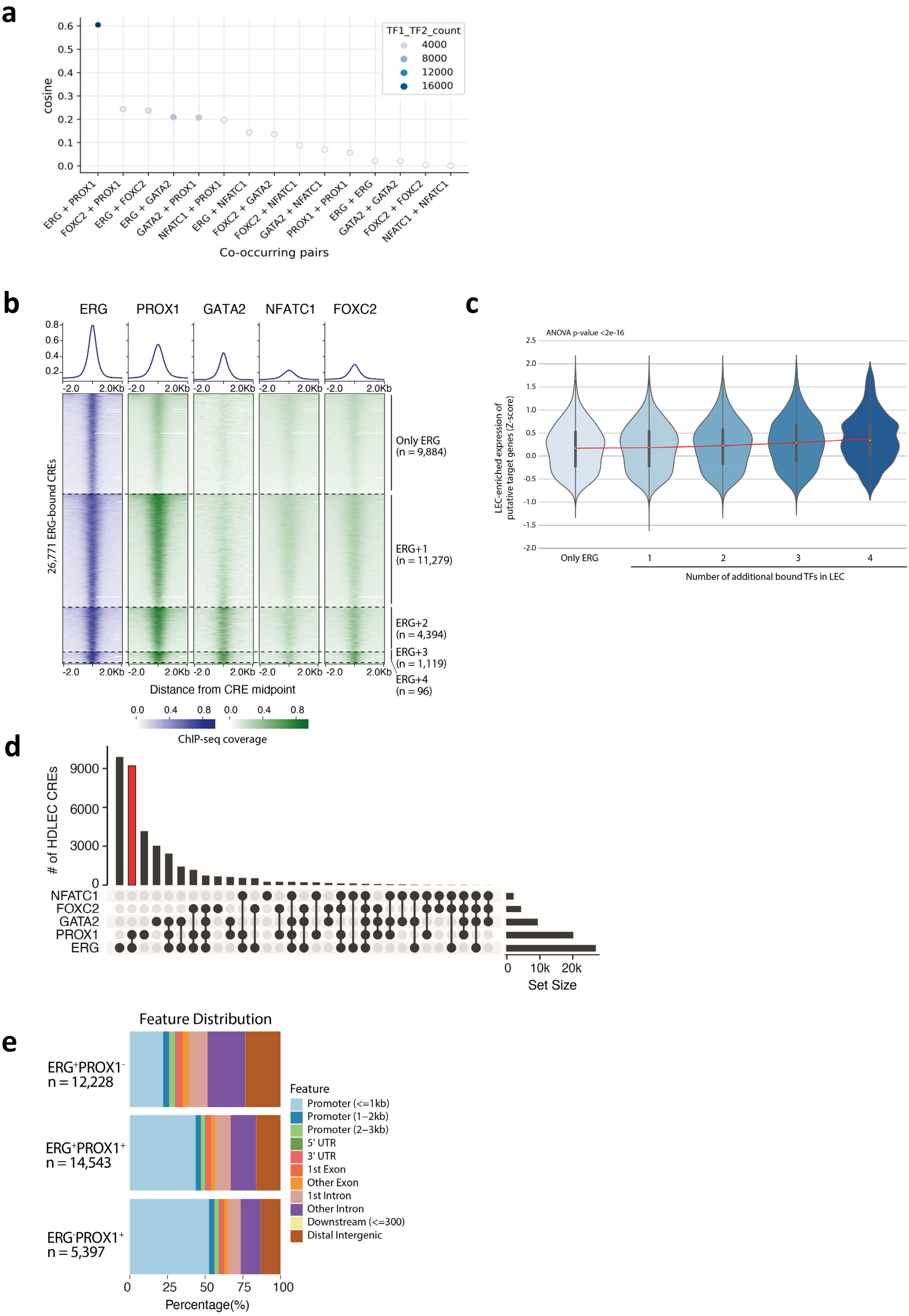

### Extended Data Figure 6, continued

f

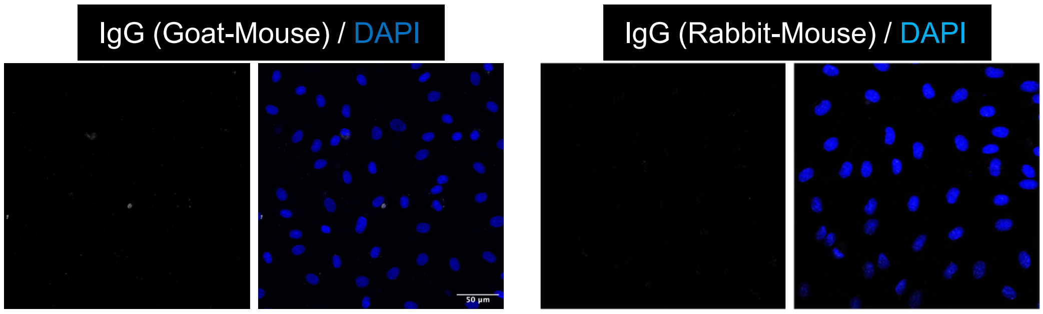


g

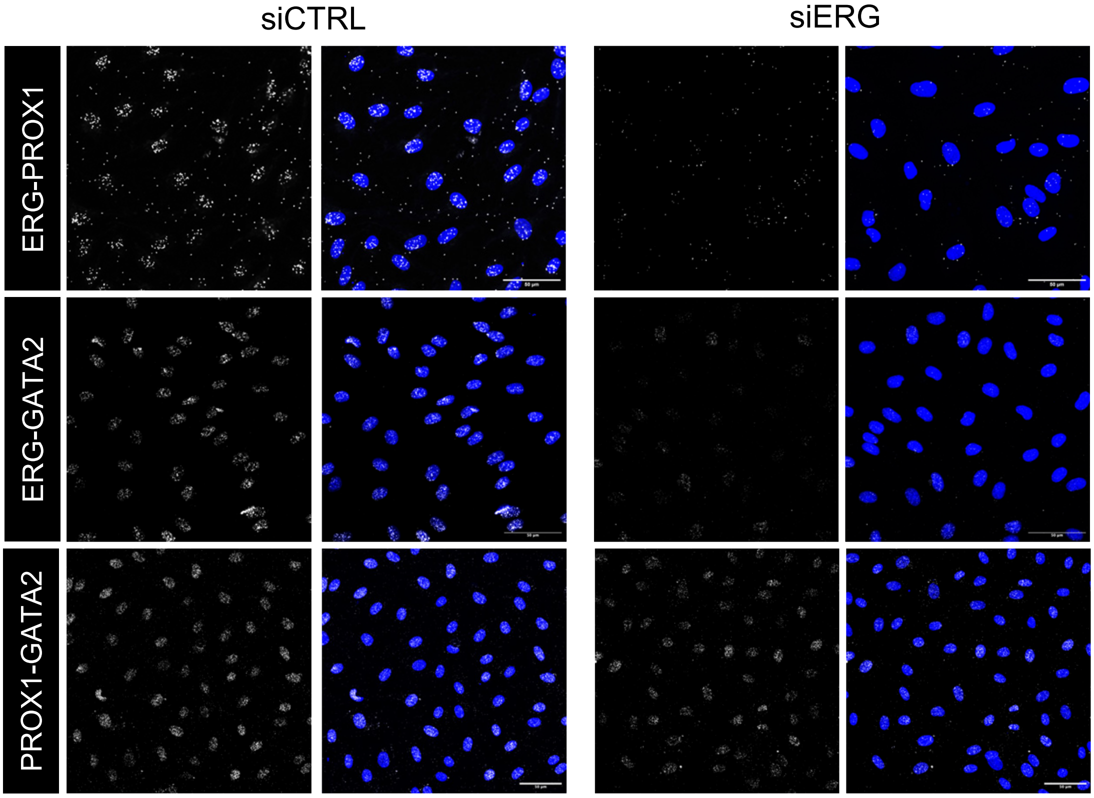


h

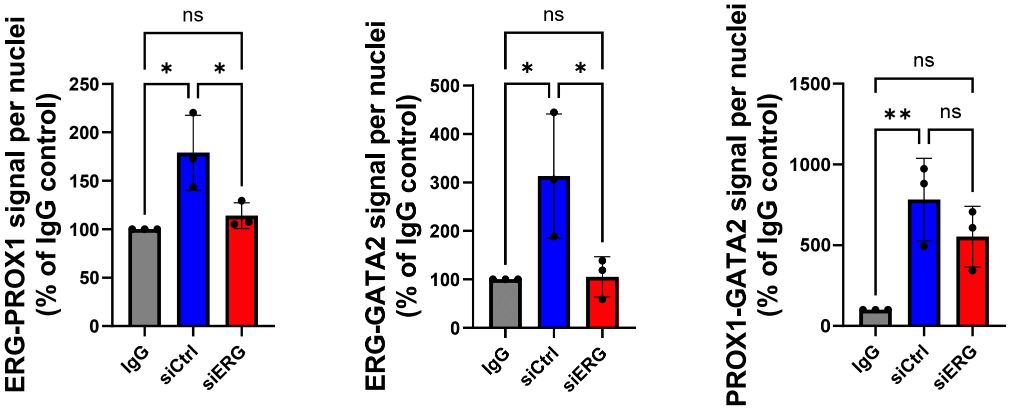

**Extended Data Figure 6.** Global overlap of HDLEC transcription factor binding. **a)** TF-COMB bubble plot showing the cosine scores of associations for TF pairs. Global overlap of TF binding was explored using an expanded list of ChIP-seq peaks for increased sensitivity (macs2 threshold of q-value < 0.05). Dots are coloured by the total number of HDLEC CREs intersecting both TFs. The ERG-PROX1 cosine score of 0.60 corresponds to a Z-score of 106.5. **b)** Heatmap of BPM-normalised ChIP-seq coverage (n = 1) at HDLEC CREs with evidence of ERG binding. Coverage is shown from the CRE midpoint +/-2kb. The colours correspond to the study: blue = this study and green = Kazenwadel *et al*.[42]. CREs are grouped according to those which bind 1, 2, 3 or 4 additional TFs (PROX1, GATA2, NFATC1 or FOXC2) in any combination. **c)** Violin plot of Z-scores representing gene expression enrichment in HDLEC compared with other endothelial cell types for putative target genes of HDLEC-CREs bound by ERG only, or those bound by ERG plus 1, 2, 3 or 4 additional TFs. **d)** UpSet plot showing the intersection of HDLEC CREs with ChIP-seq peaks for ERG, PROX1, GATA2, FOXC2 and NFATC1. The set size represents the total number of CREs which intersect with a ChIP-seq peak by at least 1bp. The intersection of ERG and PROX1 is highlighted in red. **e)** Functional annotation for the different classes of HDLEC CRE, grouped by ERG and/or PROX1 enrichment. **f)** Representative images of proximity ligation assay (PLA) in HDLEC using selected IgG as negative controls corresponding to the host species of the primary antibodies used. **g)** PLA showing nuclear interactions for ERG-PROX1 (top), ERG-GATA2 (middle), and PROX1-GATA2 (bottom) in HLDEC transfected with non-specific siRNA (siCtrl) or siRNA to ERG (siERG) for 48 hr. Nuclei are identified by DAPI (blue). Scale bar, 50 µm. **h)** Quantification PLA signal intensity per nuclei, expressed as a % of the signal intensity in relevant IgG control. One-way ANOVA, n=3, ns = not significant, * P < 0.05, ** P < 0.01.

### Extended Data Tables

| Family | Variant (NM_182918.4) | Genomic coordinates (GRCh38) | Exon | CADD | REVEL | SpliceAI | GnomAD v4.1 | ClinVar |
|---|---|---|---|---|---|---|---|---|
| 1[a] | c.671_672del p.(Thr224ArgfsTer15) | chr21:38402557 CTG>C | 5/10 | 35 | NA | NA | Absent | Pathogenic (VCV002637894.2) |
| 2 | c.1163A>G p.(Tyr388Cys) | chr21:38383680 T>C | 10/10 | 31 | 0.9 | 0 | Absent | - |
| 3 | c.917C>T p.(Pro306Leu) | chr21:38390997 G>A | 9/10 | 34 | 0.39 | 0.01 | Absent | - |
| 4[a] | c.1386del p.(Asn463ThrfsTer42) | chr21:38383456 TG>T | 10/10 | 16.03 | NA | NA | Absent | - |
| 5[a] | c.543del p.(Ser182AlafsTer22) | chr21:38403554 TG>T | 4/10 | 23.6 | NA | NA | 1 heterozygous | Pathogenic (VCV002637921.1) |
| 6[a] | c.1338dup p.(Ala447CysfsTer19) | chr21:38383504 C>CA | 10/10 | 34 | NA | NA | Absent | - |

**Extended Data Table 1.** Annotation of the six *ERG* variants identified in this study. Genomic coordinates (GRCh38), nucleotide and predicted protein changes are summarised for canonical endothelial ERG isoform NM_182918.4. Variant pathogenicity predicted by CADD, REVEL, and SpliceAI. [a]Reported by Greene et al., 2023.

| H-bond | No Mut. mean (%) | No Mut. std | Mut. mean (%) | Mut. std | Delta | Pvalue |
|---|---|---|---|---|---|---|
| 340-333 | 59.147 | 3.550 | 50.849 | 3.790 | -8.298 | 0.007 |
| 337-335 | 11.646 | 11.610 | 0.112 | 0.067 | -11.534 | 0.090 |
| 384-338 | 52.002 | 19.005 | 73.827 | 3.041 | 21.825 | 0.061 |
| 386-341 | 55.854 | 3.585 | 47.856 | 1.597 | -7.998 | 0.005 |
| 357-352 | 68.746 | 5.000 | 75.640 | 2.497 | 6.894 | 0.034 |
| 364-359 | 25.840 | 5.143 | 20.168 | 1.389 | -5.671 | 0.068 |
| 388-362 | 27.474 | 4.385 | 0.000 | 0.000 | -27.474 | 0.000 |
| 369-365 | 49.811 | 1.798 | 61.133 | 2.287 | 11.322 | 0.000 |
| 373-369 | 22.661 | 4.030 | 44.503 | 10.243 | 21.842 | 0.006 |
| 374-370 | 46.771 | 1.895 | 75.819 | 7.892 | 29.048 | 0.001 |
| 377-372 | 46.538 | 2.110 | 38.920 | 7.949 | -7.618 | 0.099 |
| 391-377 | 65.216 | 7.148 | 54.049 | 9.897 | -11.167 | 0.079 |
| 391-379 | 25.970 | 16.387 | 2.186 | 2.008 | -23.783 | 0.031 |
| 387-381 | 44.588 | 2.626 | 18.705 | 3.987 | -25.883 | 0.000 |
| 391-389 | 23.463 | 15.651 | 2.731 | 2.964 | -20.732 | 0.040 |
| 397-394 | 7.741 | 3.168 | 12.829 | 4.516 | 5.088 | 0.077 |

**Extended Data Table 2.** Hydrogen bond frequencies between ERG residues in the ETS domain that are significantly affected in the *ERG* p.Y388C variant.

| Family | ID | HGVSp | Sex | Lower limb lymphoedema (Age of onset) | Upper limb lymphoedema (Age of onset) | Genital oedema | Systemic involvement | Ig and albumin | Recurrent cellulitis | Venous incompetence | Cardiovascular problems | Lentigines | Other |
|---|---|---|---|---|---|---|---|---|---|---|---|---|---|
| 1 | II.1 | p.Thr224ArgfsTer15 | M | Y (<1y) | N | Y | Y | Low | Y | Y | | Few | Persistent warts |
| 2 | II.1 | p.Tyr388Cys | F | L (<10y), R (21-25y) | L (41-45y) | N | N | Normal | N | Y | | Multiple | |
| 2 | I.1 | p.Tyr388Cys | F | Y (>50y) | N | N | N | | N | Y | Y | | |
| 4 | II.1 | p.Asn463ThrfsTer42 | F | Y (teens) | Y? | | | | Y | | Y (26-30y) | | Obesity; hyperlipidaemia |
| 4 | II.2 | p.Asn463ThrfsTer42 | M | Y | | | | | | | Y | | Obesity; seizures |
| 5 | II.2 | p.Ser182AlafsTer22 | F | Y | Y | | N | | Y | | Y | | Macrocephaly; renal malrotation; possibly obesity |
| 5 | I.2 | p.Ser182AlafsTer22 | M | N | N | | N | | | Y | Y | | Hyperlipidaemia; possibly obesity; macrocephaly |

**Extended Data Table 3.** Clinical summary of primary lymphatic anomaly patients with confirmed *ERG* variants
